# Use of animal-derived products for medicinal and belief-based purposes in urban cities of southwestern Nigeria: a One Health perspective

**DOI:** 10.1101/2025.10.29.25339033

**Authors:** SN Akpan, R Buij, F van Langevelde, LF Thomas, P van Hooft, EAJ Cook, DM Zimmerman, JM Hassell, SP Masudi, CT Happi, AN Happi

## Abstract

The use of animal-derived products for medicinal and spiritual purposes, also called zootherapy, is a significant component of traditional medicine in many parts of the world. However, little is known about the dynamics and impact of these practices, especially in developing urban cities. This study investigated the therapeutic and belief-based use of wildlife in urban cities in Southwest Nigeria, assessing its potential implications for One Health. A mixed methods cross-sectional study design incorporating semistructured questionnaires (n=31), focus group discussions (n=4) and participants observations was used to gain insights into the socio-demography, animal species, practices, and knowledge levels of practitioners. Also, we conducted a literature search of databases (Google scholar, PubMed) for data on zoonotic pathogens associated with the animal species. Data was analysed by descriptive statistics and thematic analyses. The results revealed a total of 49 practices involving 41 animal species, with family heritage as the main source of practitioners’ knowledge acquisition. Overall, 44% (18/41) of the animal species were classified as vulnerable, near-threatened, endangered or critically endangered on the IUCN Red List, and 95% (39/41) belonged to taxa documented as zoonotic pathogen vectors, reservoirs or hosts. The use of wildlife-derived products for medicinal and belief-based purposes constitutes a potential source of zoonotic spillover, fauna decline, and ecological imbalance. There is a need for intensified risk communication and community awareness (RCCE) programs, especially targeting the practitioners and product traders. Effective monitoring, legal enforcement, and adoption of sustainable alternatives can aid in mitigating the negative impacts of these practices in Nigeria and beyond.

## 1.0 INTRODUCTION

Traditional medicine remains a primary healthcare source for many communities in developing countries, where accessibility to modern healthcare is often limited (Akunna *et al.,* 2023). Zootherapy, the use of animal-derived products for medicinal and spiritual purposes, plays a crucial role in indigenous healthcare systems across Africa, Asia, and Latin America (Soewu, 2012; Gurumyen *et al.,* 2020). It is deeply rooted in cultural and spiritual traditions (Liu *et al.,* 2024; Patrick and Singkam, 2024). In many parts of the world, animal-based treatments are used either independently or alongside plant-based treatments to address various physical and mental health conditions (Kuralkar and Kuralkar, 2021; Ahmad *et al.,* 2023; Lemhadri *et al.,* 2024). Zootherapy practices in Africa, Latin America, and Asia involve mammals, reptiles, birds, and insects (Nieman *et al.,* 2019; Friant *et al.,* 2022). Specific animal parts, such as bones, skins, organs, and secretions, are believed to have medicinal, spiritual, or symbolic properties (Inatimi *et al.,* 2022). For example, snake fat is used for its perceived anti-inflammatory effects, whereas crocodile skin is used in dermatological treatments (Mukherjee *et al.,* 2017; Mishra *et al.,* 2020). In China and Brazil, pangolin scales and jaguar fat are similarly used for traditional healing (Alves and Rosa, 2005; Tan *et al.,* 2021).

However, the hunting and killing of wild animals for medicinal and belief-based use poses threats to wildlife conservation and public health. This could drive wildlife species to extinction. In addition, the use of wild animals in traditional medicine has been linked to emerging infectious zoonotic diseases (Lee *et al*., 2020; Mitchell, 2023; Asaaga *et al*., 2024; Fourchault *et al*., 2025). The processing and use of animal-derived products, often without proper hygiene or regulation, increases the risk of zoonotic spillover (Akpan *et al*., 2025b; Alves and Rosa, 2005; Friant *et al.,* 2022; Tumelty *et al.,* 2023). In China, for example, the illegal trade of wildlife for medicinal purposes has been implicated in the emergence of SARS-CoV-2 (Zhou *et al*., 2020). Likewise, in parts of Central Africa, it has been associated with the spillover of the Ebola virus (Magouras *et al*., 2020). The selection of animals for zootherapy is predominantly influenced by cultural beliefs, attributing healing properties to animal species believed to be associated with strength, endurance, or wisdom (Kendie *et al.,* 2018; Tan *et al.,* 2021). Preparation methods, such as drying, grinding, and mixing with herbs, are widely used (Liu *et al.,* 2024).

The potency of zootherapeutic healthcare remains a subject of debate. While practitioners may believe in its efficacy, very few studies support these practices (e.g., Al-Yousef, 2012; Alhaidar, 2012). Although such studies are generally published in peer-reviewed complementary and alternative (CAM) medicine journals, they remain mostly unverified and may even be misleading (Schmidt *et al*., 2001). Additionally, rigorous clinical trials and evaluations have largely not been conducted to assess various zootherapy potency claims (Alves and Alves, 2011). Alternative medicine enthusiasts contend that scientific investigations cannot be applied to alternative medicine because the therapeutic approach is holistic and cannot be assessed via reductionistic science, the effects are too subtle to be measured, and the treatments must be customized for each patient, making them ineligible for clinical trials (Ernst, 2008).

Southwest Nigeria, a densely populated multicultural region in sub-Saharan Africa, is a wildlife trade hub for meat and zootherapeutic practices (Akpan *et al.,* 2025a; Soewu, 2012). Awoyemi *et al*. (2022) reported that “*Yoruba*” communities in southwestern Nigeria have practiced belief-based use of wildlife-derived products for many generations. Traditional healers, known locally as *"Babalawos"* (fathers of secrets), *"Iyalawos"* (mothers of secrets), or *“Ifá”* practitioners, play a central role in these traditions (Tijani, 2024). While studying the wild meat value chain in Lagos (Akpan *et al*, 2025a), we found that the medicinal and belief-based use of wildlife products in urban city settings may be a risk factor driving wildlife exploitation, trade and health risks. Hence, additional focus is needed on this subject as a distinct yet overlapping aspect of the wild meat trade in Lagos and other developing urban cities. Many studies conducted on this topic have been fragmented, focusing either on public health, anthropology, or on ecology and conservation (Gurumyen *et al*., 2020; Friant *et al*., 2022; Green *et al*., 2022; Coals *et al*., 2024; Lemhadri *et al*., 2024; Adebowale *et al*., 2025; Fourchault *et al*., 2025) . Therefore, this study examines the implications of medicinal and belief-based wildlife uses through the holistic lens of One Health, which encompasses human, animal, and environmental health, drawing parallels to similar practices in other regions of the world. The findings from this study contribute to a broader global understanding of the intersection between traditional medicine, biodiversity conservation, and public health.

## 2.0 METHODS

### 2.1. Ethical Approval

This study received ethical approval from the National Human Research Ethics Committee of Nigeria (NHREC), with approval number NHREC/01/01/2007-07/12/2022.

### 2.2. Data collection

A mixed methods study approach was employed, incorporating questionnaire administration and focus group discussions (FGDs). Expanding on a previous study that mapped the wild meat value chain in Lagos metropolis (Akpan *et al*., 2025a), the research team visited wildlife product sections of open markets and traditional healing homes across Lagos and four other states in southwestern Nigeria: Ogun, Oyo, Ondo and Osun (Figure 1). The specific towns visited were Surulere, Ikeja, Epe, Abeokuta, Ijebu-ode, Ibadan, Oyo town, Akure, Ondo town, Osogbo, and Ilesha. Figure 1 shows the map of Nigeria with the study locations.

**Figure 1.**
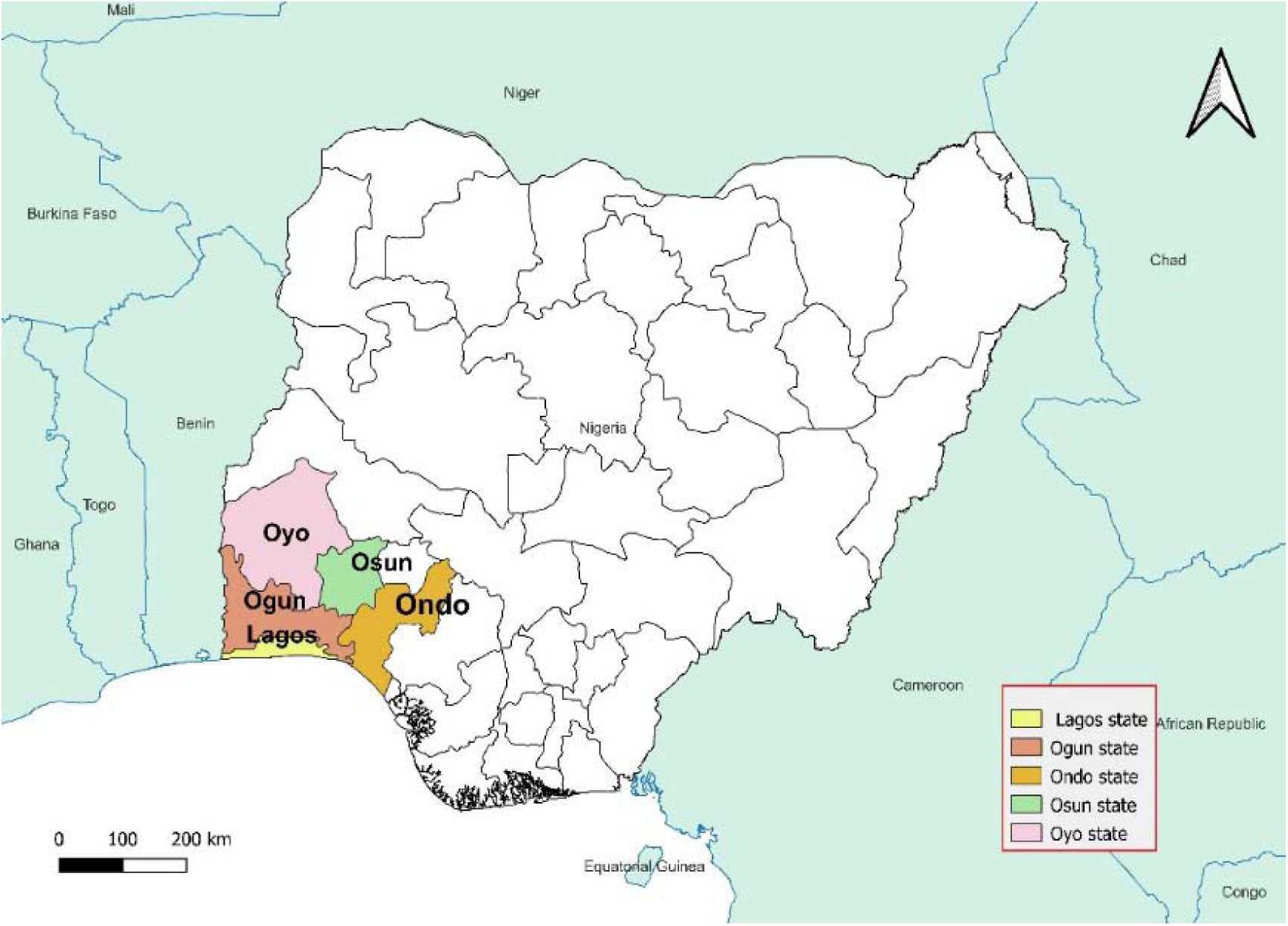
Map of Nigeria showing the study locations

Using a local language interpreter, a purposive sampling approach was used to randomly select study participants, with a focus on individuals involved in the trade or use of wild animal parts for traditional healthcare and other belief-based uses. Using a project information sheet (**S1**), we described the project objectives, and administered semistructured questionnaires (**S2**) to thirty-one (n=31) study participants who gave consent (S3). Additionally, following a guide (**S4**), four FGDs (one per state) were conducted with practitioners who declined consent to complete questionnaires but preferred to speak in groups. All the participants gave verbal consent, as witnessed by the lead author. Additionally, the participants’ observations were recorded, and photos were taken of the wildlife body parts used for zootherapy, as seen by the researchers during the field data collection process.

### 2.3. Data analysis

Species seen or photographed during field observation were phenotypically identified by experts in the research team, and the categorical data were analysed via descriptive statistics. Qualitative data were analysed via thematic analysis. Following the methods described by Braun & Clarke (2006), we employed a deductive-inductive thematic approach to analyse the FGD. We read through the transcripts repeatedly, identifying recurring or significant patterns of meaning and coding them to develop detailed accounts of participants’ knowledge and experiences. We then grouped the codes to reveal subthemes, iterating to ensure the groupings were appropriate, and that the emergent themes transcended individual experiences and accounts. To enhance a better understanding of zoonotic risks, we conducted a literature search of two databases (Google Scholar and PubMed) for information on zoonotic pathogens that were associated with the wildlife species reported in our study. We combined free-text keywords and controlled vocabulary (e.g., MeSH terms in PubMed) using Boolean operators as follows: (“zoonoses”[MeSH Term] OR “zoonotic diseases" OR “spillover”) AND (“pathogens” OR “viruses” OR “bacteria” OR “parasites”) AND (“wildlife” OR “wildlife species” OR “wild animals”) AND (“zootherapy” OR “medicinal use” OR “ethnomedicine”). The search covered studies that were published between January 2000 to October 2025.

## 3.0 RESULTS AND DISCUSSION

### 3.1 Socio-demography

The results revealed an equal gender balance among the respondents (16 males, 15 females). Approximately half of the respondents (15/31) were between 51 and 65 years old, followed by 10 respondents (10/31) in the 66–80 age group. Most respondents (18/31) had 20--29 years of experience, followed by 30--39 years (5/31) and 0--9 years (4/31), respectively. Traditionalists constituted the largest group of practitioners (15/31), with primary school emerging as the most predominant level of education of the respondents (16/31), followed by informal education (10/31) and secondary school (5/31). This shown in Table 1.

**Table 1:** Socio-demography of the questionnaire respondents

| Category | Religion | Frequency (%) |
| --- | --- | --- |
| **Gender** | Male | 16 (51.6) |
|  | Female | 15 (48.4) |
| **Age range** | 21-35 | 2 (6.5) |
|  | 36-50 | 4 (12.9) |
|  | 51-65 | 15 (48.4) |
|  | 66-80 | 10 (32.3) |
| **Religion** | Traditionalist | 15 (48.4) |
|  | Muslim | 9 (29.0) |
|  | Christian | 5 (16.1) |
|  | Other | 2 (6.5.) |
| **Experience** | 0-9 | 4 (12.9) |
|  | 10-19 | 3 (9.7) |
|  | 20-29 | 18 (58.1) |
|  | 30-39 | 5 (16.1) |
|  | 40-49 | 1 (3.2) |
| **Education** | Primary school | 16 (51.6) |
|  | Secondary school | 5 (16.1) |
|  | Tertiary education | 0 (0) |
|  | Informal education | 10 (32.3) |
|  | None | 0 (0) |

The level of education also plays a role in people’s knowledge of environmental, animal, and human health risks. Education equips an individual with knowledge, attitudes, and skills to recognize risks and prevent harm to him or herself, his or her environment, and the public (Hahn & Truman, 2015). Hence, the low level of education observed in this study suggests that practitioners may have engaged in risky practices, at least partly due to insufficient knowledge and awareness, as alluded to by Masudi *et al*. (2025). Similarly, religious affiliation plays a crucial role in shaping the perception and practice of zootherapy (Ahmad *et al*., 2023). Hence, the involvement of practitioners with different religious beliefs suggests that there may be variations in zootherapy application on the basis of religious doctrines (Ademiluka *et al*., 2023). Additionally, the high degree of involvement of traditionalists in zootherapy, as seen in this study, suggests a strong connection between these practices and African traditional religious belief systems. The predominance of participants older than 50 years (25/31) in this study is consistent with previous studies reporting that traditional zootherapy and belief-based use of wildlife were practiced primarily among older individuals (Akpa-Inyang and Chima, 2023; Ahmad *et al*., 2023). We posit that the low level of involvement among younger persons may be due to increased knowledge of infectious disease concerns related to zootherapy or a gap in intergenerational knowledge transfer. This gap is often attributed to factors such as globalization and urbanization, which influence younger individuals to adopt modern lifestyles, distancing them from traditional practices (Nanda, 2023).

Thematic analysis of the FGDs revealed seven themes: (i) species and practices, (ii) hygiene and safety, (iii) urban preference, (iv) importation (v) wild meat as a modulator, (vi) knowledge and beliefs, and (vii) sustainability and conservation. Table 2 presents the overarching themes, subthemes, and their descriptions.

**Table 2:** Code book showing themes, subthemes, and their descriptions

| Themes | Codes<br>(subthemes) | Description |
| --- | --- | --- |
|  | Species | For comments on the wildlife species used by study participants |
| Species & Practices | Uses | For comments describing practices and the utilization of wildlife-derived products |
| Hygiene & Safety | Products hygiene | Used to capture comments on hygiene measures taken by participants |
|  | Products safety | Used to capture comments on the safety of wildlife-derived products |
| Importation | Products scarcity | For comments on the scarcity of local species or their products |
|  | Products sourcing | Captures comments on sourcing of products internationally |
| Urban preference | Income | Captures comments on the financial motivations of participants |
|  | Accessibility | Captures comments on participants' ease of access to clients, and clients' ease of access to their services |
| Wild meat as a modulator | Demand | Used to capture comments on the local demand and supply of wildlife-derived products |
|  | Wild meat trade | Used to capture comments on wild meat hunting and trade |
| Knowledge & belief systems | Zoonoses | Used to capture comments on how wildlife-human disease transmission risks |
|  | Culture | Captures comments on cultural beliefs |
| Sustainability & Conservation | Wildlife resilience | Captures comments on the high fecundity of wildlife species |
|  | Wildlife decline | Captures comments on wildlife population decline |

### 3.2 Species and practices

The study participants reported a total of forty-nine (49) practices, cutting across thirty-eight (41) wildlife species. Some participants, however, declined to comment on the use of four species and three products, citing the need to keep some practices discreet. The reported species, their products, associated zoonotic pathogens, and their conservation statuses are shown in Table 3.

**Table 3:**
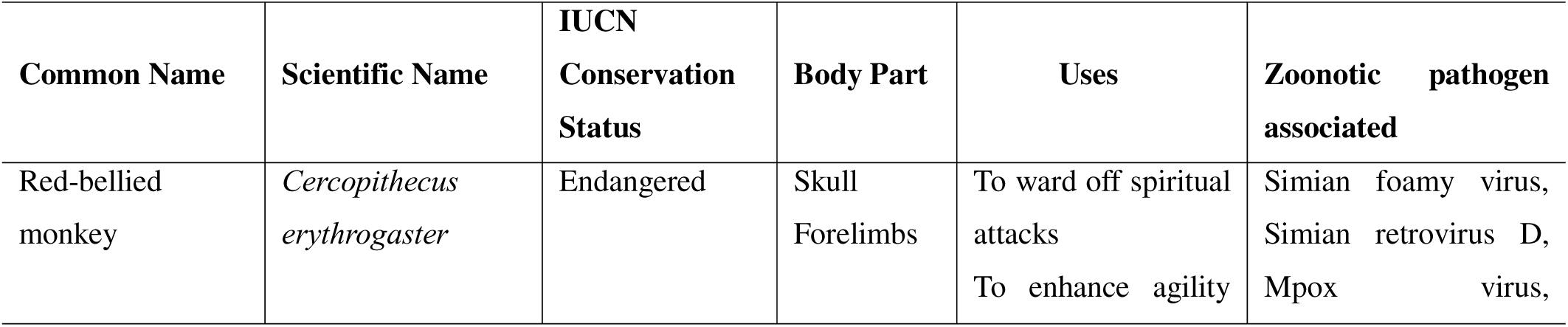
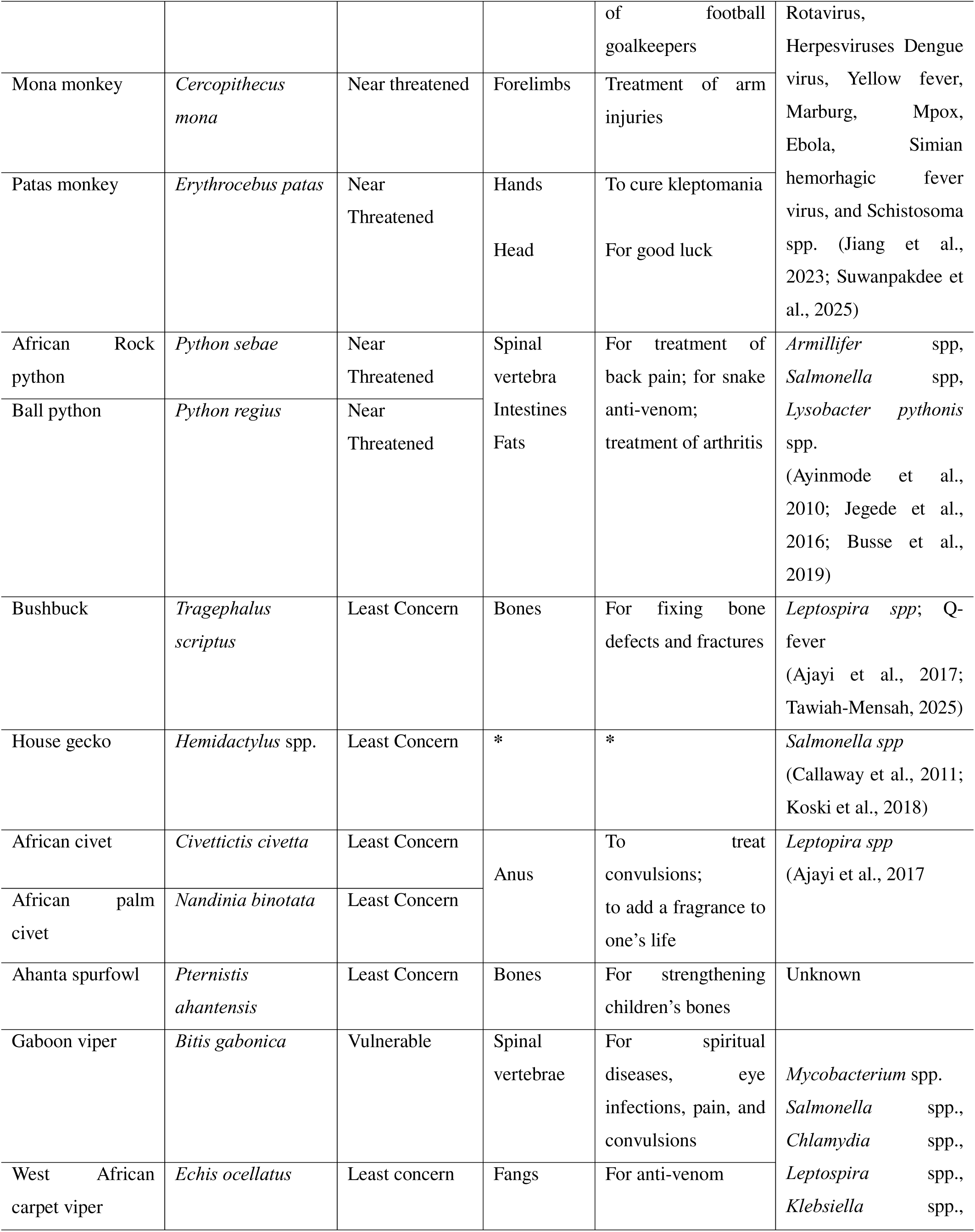
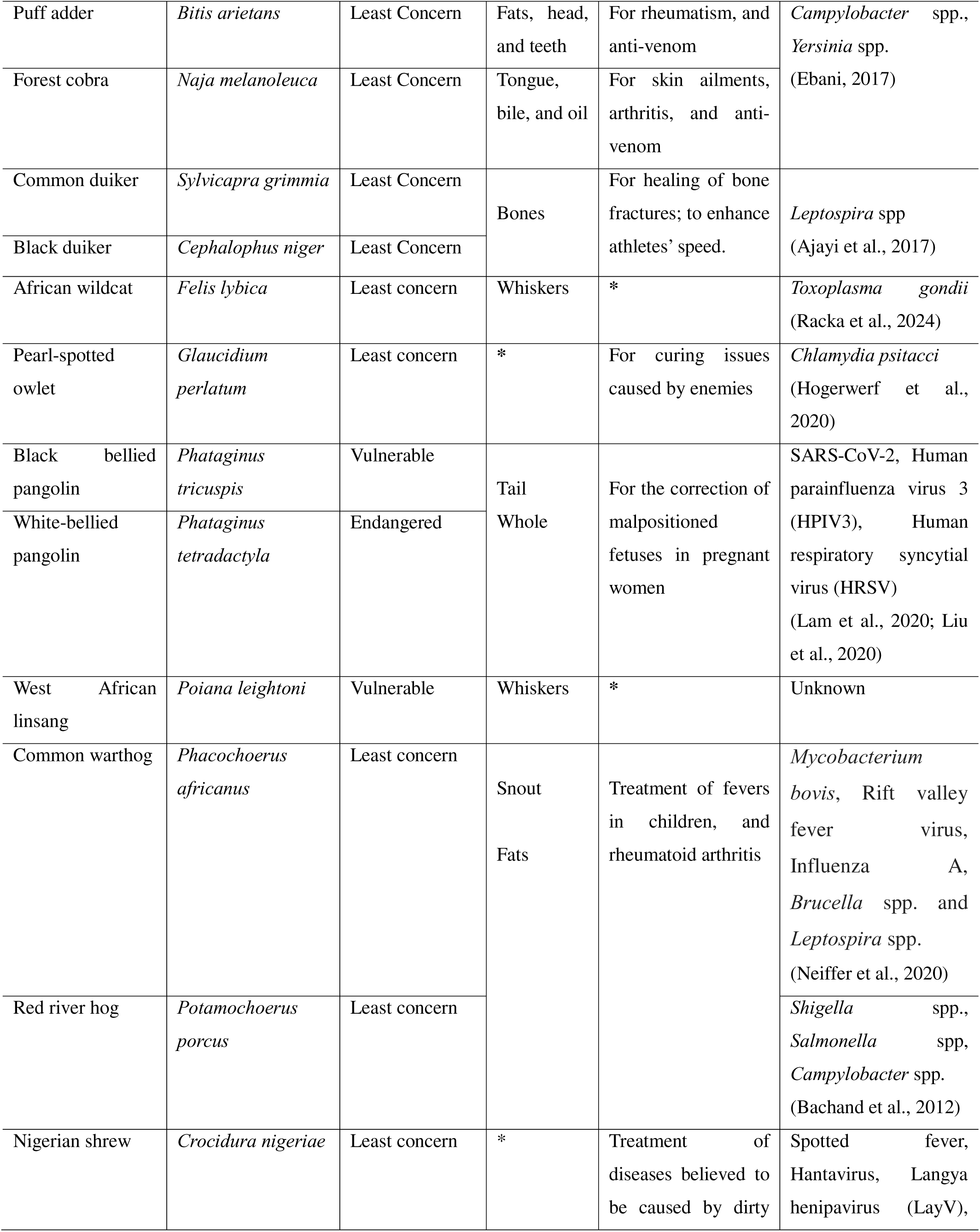
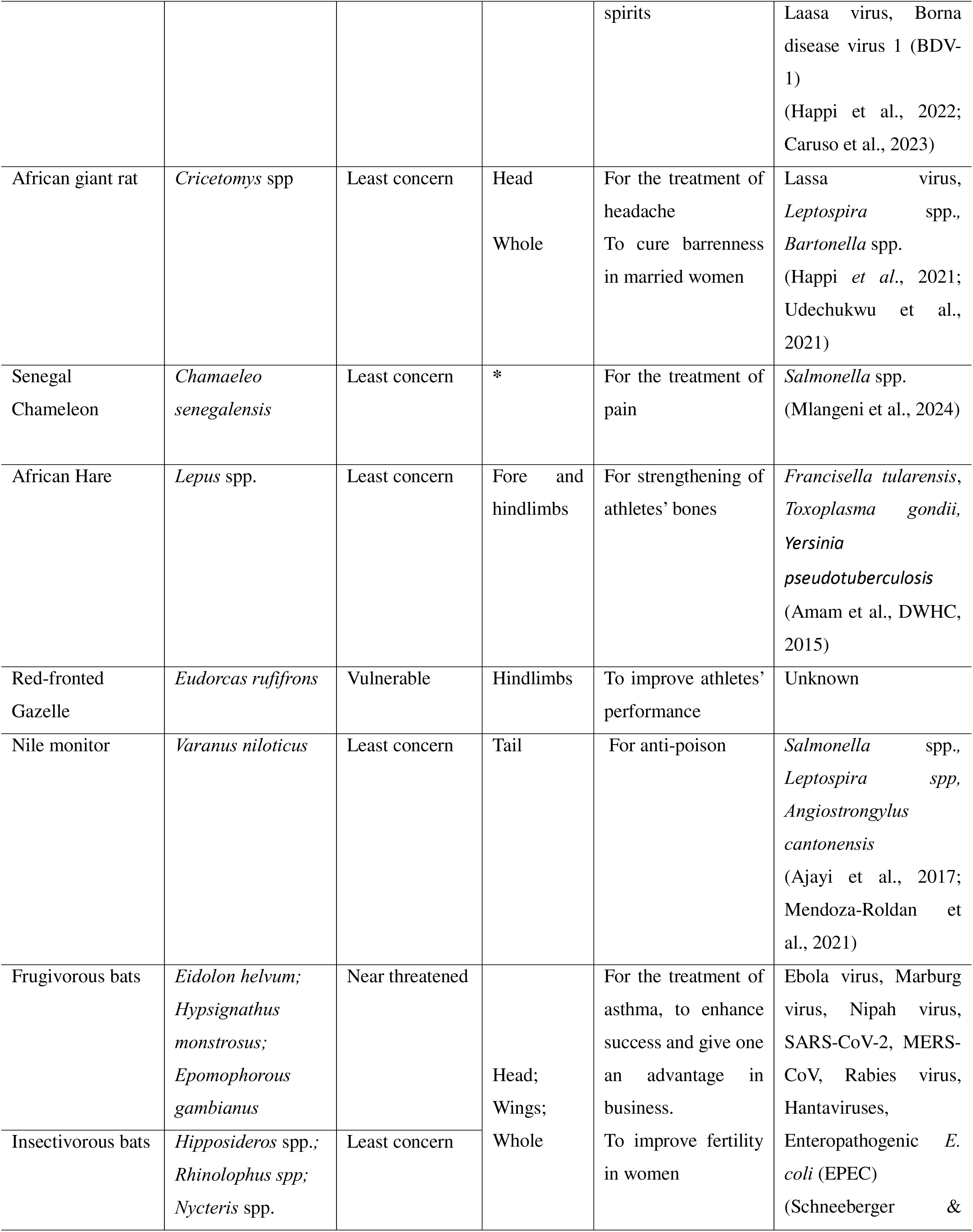
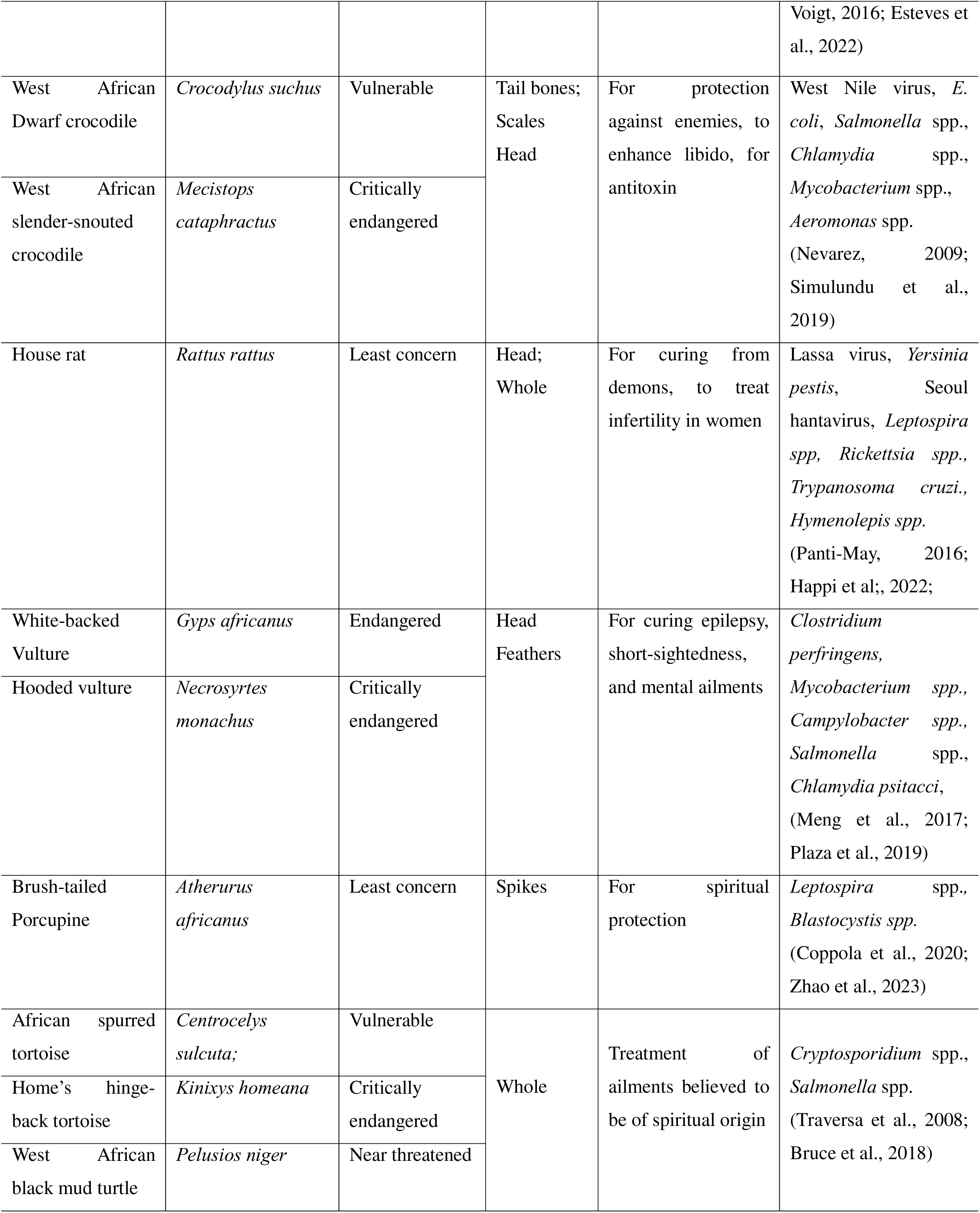
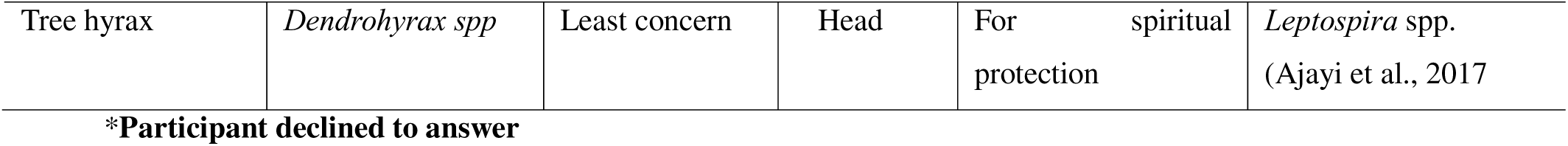
Wildlife species, conservation statuses, derived products, uses, and associated zoonotic risks

These were (i) forest/jungle species: red-bellied monkeys, mona monkeys, patas monkeys, blue duikers, sitatungas, red-fronted gazelles, black-bellied pangolins, tree hyraxes, white-bellied pangolins, forest cobra, Nigerian shrews, giant-pouched rats, house rats, common warthogs, red river hogs, civets, and Ahanta spurfowls; (ii) savanna/grassland species: African hares and brush-tailed porcupines; (iii) wetland species: dwarf crocodiles, slender-snouted crocodiles, and mud turtles; (iv) desert species: monitor lizards; and (v) general species: Ahanta spurfowls, rock pythons, ball pythons, puff adders, spotted linsangs, Gaboon vipers, chameleons, bats, house geckos, owlets, vultures, and tortoises. In terms of specific practices, our study revealed that the whole heads of Nigerian shrews and Gambian giant-pouched rats are believed to have the ability to cure demon-related illnesses and headaches. Hare and gazelle limbs are believed to enhance athletic performance, whereas crocodile bones are thought to counteract poisons. Vulture heads are used to treat epilepsy, porcupine spikes are believed to offer spiritual protection, and tortoises are used for spiritual healing, while some species’ applications remain undisclosed (Table 3).

The respondents also reported that monkey bones and ligaments (Figure 1) were used to treat arm injuries in football goalkeepers, whereas python spinal vertebrae, intestines and fat aid in backache, arthritis and antivenom production. The bones of Ahanta spurfowl (popularly referred to as “bush fowl” by the locals) were used to strengthen children’s bones, whereas the Gaboon viper vertebrae were used for the treatment of diseases believed to be of spiritual origin. Bile, tongue, and fat derived from forest cobras contribute to antivenom and skin remedies. Pangolin scales were used to correct fetal malposition, and bush-pig snouts were used to treat fever in infants. Figure 2 shows photos of some wildlife-derived products photographed during the field data collection.

**Figure 2.**
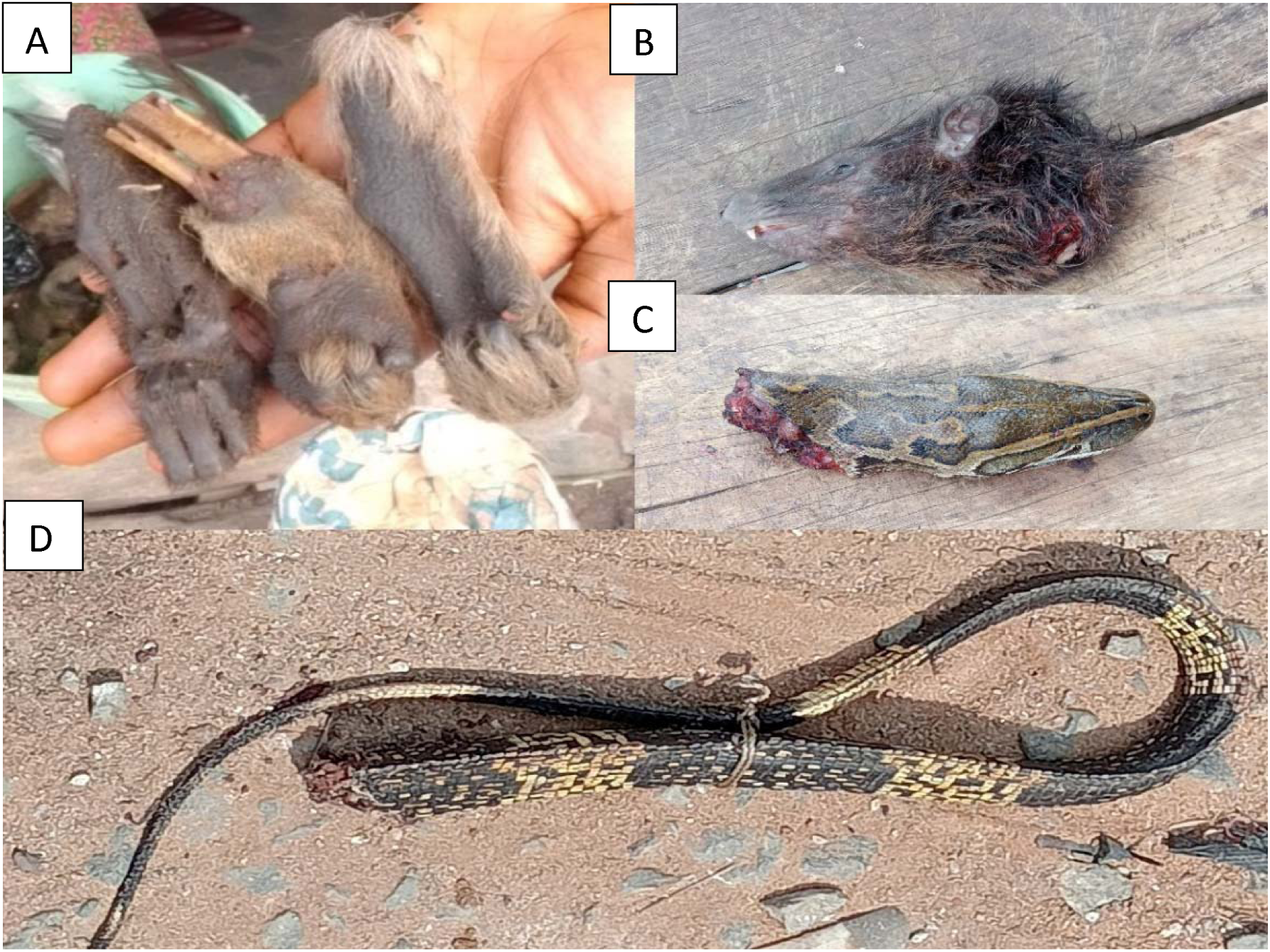
Some wildlife parts used for medicinal and belief-based purposes across the study sites:[A] Preserved forelimbs of monkeys of different species [B] Whole head of a western tree hyrax [C] Whole head of an African rock python [D] Tail of a Nile monitor lizard

The reliance on wildlife products for these practices raises concerns regarding the overharvesting of wildlife populations. The conservation statuses of the species revealed that 44% (18/41) of the reported species were classified as vulnerable (e.g., red-fronted gazelle, black-bellied pangolin), nearly threatened (e.g., rock python, mona monkey), or endangered (e.g., white-bellied pangolin) (IUCN, 2023). Others, such as the hinge-back tortoise, slender-snouted crocodile, and hooded vulture, are even critically endangered (IUCN, 2023), necessitating stronger conservation measures across the study areas. Previous studies have demonstrated that unregulated wildlife use can drive species to extinction, as observed in the case of pangolins, which are now critically endangered because they are poached for traditional medicine and international trafficking (Challender *et al*., 2020). Without immediate conservation interventions, the depletion of these wild animal species could have cascading ecological consequences, including the loss of critical ecosystem services, such as the natural pest control services provided by pangolins and vultures (Gao *et al*., 2022; Santangeli *et al*., 2024).

### 3.3. Hygiene and safety

With respect to safety concerns related to the processing and preparation of wildlife-derived products for traditional medicines, one participant stated:

*“Wild animals are from the wild, which is pure, so most of them don’t have any germs. However, we usually heat the animal parts on fire, to kill anything [germs] that may be there”*.

Another FGD participant stated:

> “We can use the same product for different people. For example, one can use the same monkey hands [forelimb] to cure stealing addiction [kleptomania] for up to 20--30 persons. There is no issue of passing [transmitting] any germs to people by contact with the same animal part…..it is not possible”.

While the heating or burning process of wildlife products (as seen in the excerpt) may inhibit certain heat-labile zoonotic pathogens that may be present, heat treatment is not always effective. Although many foodborne pathogens (e.g., *Salmonella spp*., *E. coli*) can be killed when exposed to high temperatures of 65°C and above (Spinks *et al*., 2006), some pathogens cannot be destroyed by high temperatures. For example, the spores of *Clostridium botulinum* and *Baccillus anthracis* (causative agents of botulism and anthrax) can withstand intense heat for extensive periods of time (Clery-Barraud *et al*.,2004; Wilkins, 2014). Furthermore, given the unstandardized and unmonitored state of the practices, the heating temperature (and duration) deployed by practitioners for the disinfection of animal products is unknown. Recent disease outbreaks demonstrate the devastating health and socioeconomic consequences that can arise from zoonotic spillovers (Agusi *et al*., 2022; Tajudeen *et al*., 2022; Fourchault *et al*., 2025). The absence of standardized handling and processing procedures further increases the risk of disease spillover among traditional healers (Haq *et al*., 2024; Tran and Xie, 2024). Moreover, the lack of regulatory oversight in wildlife-based medicinal practices increases the likelihood that contaminated animal products may inadvertently be used in treatment, further increasing the risk of zoonotic transmission (Magouras *et al*., 2020; Rush *et al*., 2021).

Some practitioners use animal wastes, such as urine and feces, for human treatment (Soewu, 2008), a practice that favors the transmission of a wide variety of zoonotic pathogens. Additionally, a study conducted in Nigeria revealed the presence of antimicrobial-resistant (AMR) bacteria (*Bacillus* spp. 100%, *Staphylococcus* spp. 12.5%) in cow urine that was used for zootherapy (Ogunshe *et al*., 2010). This can cause the cross-transfer of AMR genes from the material to the human patient and the human population at large.

### 3.4 Importation

With respect to the sourcing of the products, two FGD participants reported that, owing to the unavailability of certain animal-derived products, they sourced supplies from nearby countries: Cameroon and the Republic of Benin. However, no participant in this study reported exporting products.

Excerpts:

> “…several times the animal part I need is not available in our area……an animal like a leopard, for example. Sometimes the animal may be in our area, but the hunters are not lucky enough to see it. In that case, I talk to some connections in that place after you cross from Lagos….Benin. When I send money, they send it [the products] for [to] me” – FGD participant

> *“Apart from also treating people, I also supply the products to some of our people, especially the elderly ones who cannot move around as before. When we don’t have some things here, I get the ones some traders bring from Cameroon and the Benin Republic.” –* FGD participant

International trade in wildlife products increases the risks of transmission of transboundary animal diseases (TADs) such as foot and mouth disease (FMD), African swine fever, anthrax, peste des petits ruminants (PPR), lumpy skin disease (LSD), and highly pathogenic avian influenza (HPAI) (Yadav *et al*., 2020). These diseases threaten animal health and food security and may lead to marked economic losses (Yadav *et al*., 2020). Hence, unmonitored cross-border sourcing of wildlife products (as reported in this study) complicates regulation and underscores the need for sustainable strategies to reduce transboundary disease risk and pressure on wildlife resources.

### 3.5. Urban preference

The FGD participants’ responses indicated that the practices did not differ between rural and urban areas. However, there was a growing interest of zootherapists in practicing in urban areas due to income considerations (i.e., the financial strength of clients in urban areas). This is seen in the excerpts:

> *“Some of us live in the rural villages but come here to treat people, because here there are many clients, they have the money, and they usually pay better for our services….hahahahaha [laughs]….you understand?* - FGD participants

> *“Wo o [look], many big men [wealthy persons] were coming to my town to meet me, but sometimes they complain, because it is very far and the road is bad. Because of that, with the help of my eldest daughter, I moved to this part of Lagos. Since I am here, it is very easy for clients to visit me and refer other people who need me”* - FGD participant

> *“…being in the city is good for us. Apart from making more money, the bushmeat market in the city is bigger, and one can get the attention of many hunters and get the needed raw materials.” -* FGD participant.

The excerpts show that practitioners’ preference for urban centers were also driven by the perceived societal status of clients in these areas, and the need to improve clients’ ease of access to their services. Consistent with this finding, Coals *et al*. (2024) state that wildlife products and services are traded and utilized across geographic and socioeconomic gradients, including urban cities, due to their integration into the global economy. This suggests that human wildlife product interactions and zoonotic exposure risks may be more frequent in urban areas due to socioeconomic rural urban migration.

### 3.6. Wild meat as a modulator

FGD participants reported that they or other persons had consumed meat from the same animals from which they derived products for medicinal or belief-based uses. The participants who were traditional hunters stated that the demand for wild meats and wildlife products believed to have medicinal properties often stimulated their hunting expeditions and that one cannot be separated from the other, as shown in the following excerpts:

> *“The same trader selling bushmeat is the one who will sell crocodile head to you, if you need. It is the same thing. Although some of us only deal with wildlife parts used for medicine and other spiritual work, many deal with the two [both] because it is [they are] the same thing.”* - FGD participant.

> *“When we hunt a snake for their delicious meat, the first thing we do is to cut off the head and sell it to any traditional medicine dealer, or we use it by ourselves… you cannot waste it, because apart from the meat, every part of the animal is useful for other things. Then, we sell the meat to those who want to eat.” -* FGD participant.

> *“Many hunters don’t agree to enter the bush to hunt because of one tiny animal part that we need. They ask us to wait until they get a demand for bushmeat, then they go. It is from the bushmeat that they bring to us any animal parts that we need, like monkey skull” -* FGD participant.

These excerpts highlight the interdependence and intricate web linking zootherapy with the wild meat trade and consumption. This aligns with findings from other studies by Saidu & Buij (2013) and Williams *et al*. (2014), who reported that zootherapy and the wild meat trade were integrated and interdependent (Saidu & Buij, 2013; Williams *et al*., 2014), as the same harvested animals served both purposes. While our study results did not clearly establish the extent to which zootherapy demand stimulated wild meat, our findings suggest that wild meat demand plays an important role in modulating zootherapy practices.

### 3.7. Knowledge & Belief Systems

In response to a question about the potential risks of zoonotic disease through humanDwildlife contact, an FGD participant responded:

> *“Nature does not bless and curse. That is why wild animals do not pass any disease to us or anyone. As far as we are concerned, wild animals are clean [pure], and the forest is clean [pure].”*

This excerpt shows the poor knowledge of practitioners on zoonotic risk, reflecting a global challenge where traditional medicine practitioners often lack formal education on zoonoses. Akpan *et al*. (2025b) also reported the poor knowledge of actors of zoonotic risk in the processing and trade of wildlife products. For example, bats, primates, and rodents (which feature zootherapy and belief-based practices in these study locations) are globally known reservoirs of zoonotic pathogens (Friant *et al*., 2022) and can be transmitted via direct human contact or the ingestion of their products and wastes. Therefore, the general belief in the purity of wildlife may predispose practitioners and their human patients to zoonotic infections.

Another FGD participant stated:

> *“….it is part of the tradition. Every community usually has at least one of us to solve problems….or they go to where they can find us. That is the only way to handle certain [medical, spiritual] cases among our people…..and it is very effective.”*

As seen from the excerpt, strong cultural beliefs related to zootherapeutic efficacy further drove the practices (Soewu, 2012), corroborating the findings from our previous study, which reported that wild meat and related practices were driven by the culture and traditional beliefs of the “*Yorubas*”, the predominant tribe of southwestern Nigeria (Akpan *et al*., 2025a). Traditional medicine is often intertwined with religious and mystical elements, necessitating culturally sensitive public health interventions that acknowledge these beliefs while promoting safer and more sustainable practices (Souto *et al*., 2011; Aina *et al*., 2020).

Furthermore, our study results revealed that “family heritage” was the main route through which the participants acquired their knowledge of zootherapy (Table 4).

**Table 4.**
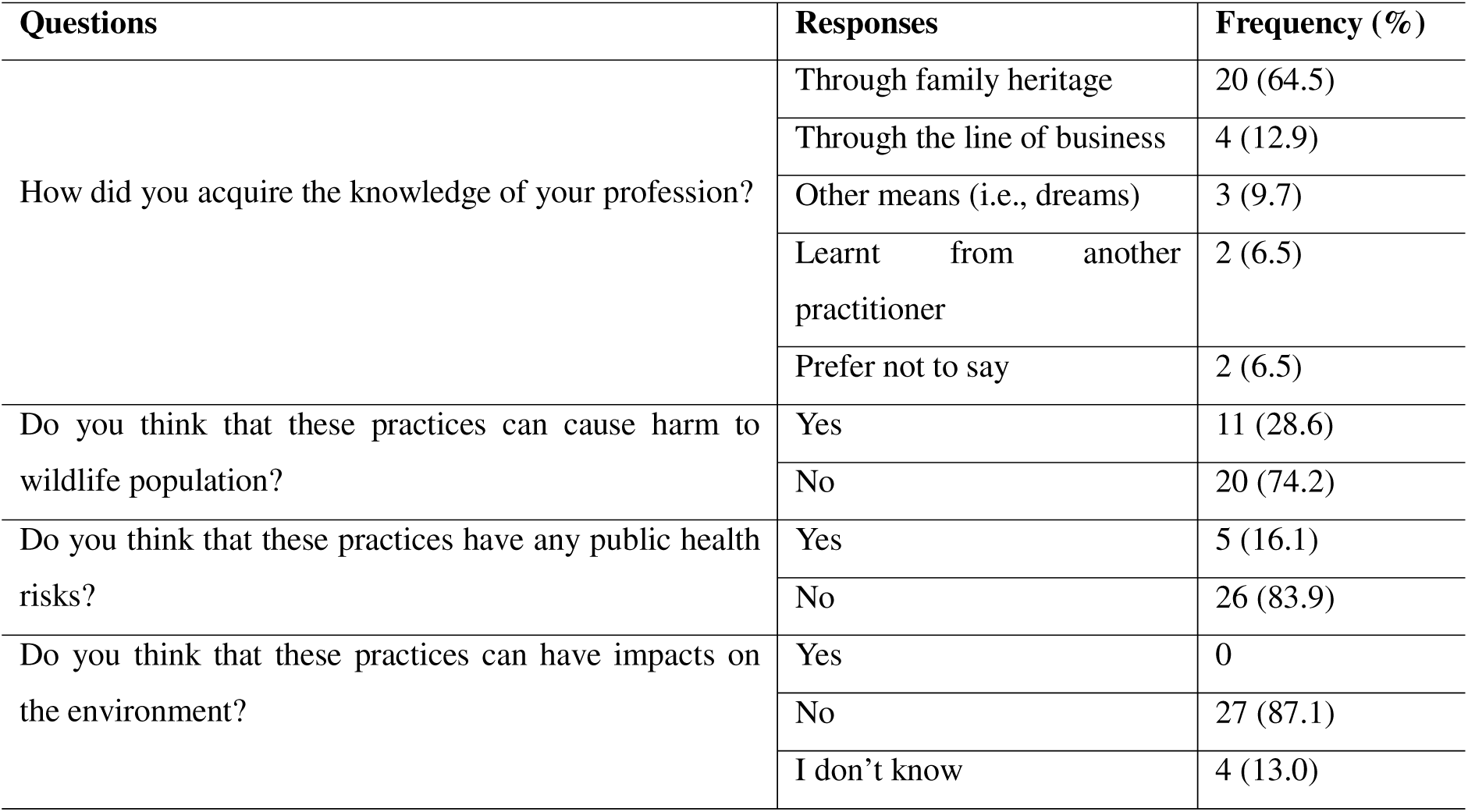
: Participant responses on knowledge acquisition, conservation, and public health risks

| Questions | Responses | Frequency (%) |
| --- | --- | --- |
| How did you acquire the knowledge of your profession? | Through family heritage | 20 (64.5) |
|  | Through the line of business | 4 (12.9) |
|  | Other means (i.e., dreams) | 3 (9.7) |
|  | Learnt from another practitioner | 2 (6.5) |
|  | Prefer not to say | 2 (6.5) |
| Do you think that these practices can cause harm to wildlife population? | Yes | 11 (28.6) |
|  | No | 20 (74.2) |
| Do you think that these practices have any public health risks? | Yes | 5 (16.1) |
|  | No | 26 (83.9) |
| Do you think that these practices can have impacts on the environment? | Yes | 0 |
|  | No | 27 (87.1) |
|  | I don't know | 4 (13.0) |

Here, children were taught through the activities of their parents and grandparents and were expected to imbibe the practices or trade as part of their ancestral inheritance. Others acquired knowledge through their line of business (4/31), whereas some reported acquiring knowledge through dreams while they slept (3/31). Only two of the respondents said they learned from other practitioners (Table 4). A triangulation of participants’ years of experience, age range, and mode of knowledge acquisition suggests that zootherapy may require extensive time to master, often through apprenticeships and intergenerational knowledge transfer. Consistent with this finding, previous studies state that traditional medicine functions as a repository of ancestral knowledge, passed down verbally through generations (Grigo, 2022; Akunna *et al.,* 2023; Rajasekharan *et al.,* 2023). However, the reliance on verbal transmission through family heritage, rather than formal documentation or scientific validation, often contributes to misinformation and unsafe practices (Soewu, 2012). A study conducted in Uganda reported that the predominance of family heritage as the primary mode of knowledge acquisition limited exposure to modern medical and conservation principles (Bagwana, 2015). As stated previously, some respondents (3/31, 9.7%) in our study reported acquiring zootherapy knowledge through dreams, further emphasizing the mystical dimensions of these practices. This phenomenon is particularly evident in shamanistic cultures, where dreams are considered conduits for spiritual guidance and the transmission of healing knowledge (Laughlin and Rock, 2014). For example, in some indigenous communities, individuals with shamanistic abilities may experience dreams that provide insights into medicinal practices, including the use of animal parts for healing purposes (Elendu, 2024). While such practices may be culturally significant, they lack scientific validation, potentially leading to misinformation and unsafe health applications (Coals *et al*., 2022).

### 3.8. Sustainability and Conservation

The majority of the respondents (20/31) in this study believed that their practices did not cause any harm to or decline in wildlife populations (Table 4), indicating that zootherapy was sustainable due to the abundance of forests and wildlife in Nigeria. However, current global conservation realities suggest otherwise. Nigeria faces challenges with the overexploitation of wildlife for medicinal use. Trade in vulture parts for traditional healing drives several vulture species towards extinction, which threatens their crucial ecological role in carcass disposal and disease prevention (Awoyemi *et al*., 2022). Chameleon overharvesting for traditional medicine and rituals has resulted in population reduction, putting the species at risk of extinction in particular locations (Adebowale, 2025). Overall, the participants stated that zootherapy was sustainable due to the abundance of forests and wildlife diversity in Nigeria. However, three participants noted that although certain species (e.g., pangolins) had become scarce, they believed in the resilience of wildlife despite anthropogenic pressures. Additionally, all the FGD participants in this study said that zootherapy was more effective than modern medicine, especially for difficult-to-treat diseases, which they believed were caused by demons. The poor knowledge expressed by participants and the lack of scientific evidence to support the potency claims of zootherapy underscore the urgent need for innovative approaches to prevent people from further indulgence in these practices to mitigate the progressive overexploitation of already declining wildlife resources in Nigeria.

## 4.0 CONCLUSION

Despite increased access to modern healthcare in developing urban areas, the belief-based use of wildlife-derived products persists due to its deeply rooted cultural connections. These beliefs stem from ideologies that encourage wildlife-based solutions for human nutrition and health. Diseases are viewed as demonic possessions of humans, cured only through the use of wildlife-derived products and enhanced by spiritual incantations. The major concerns of these practices are their potential role in zoonotic disease transmission (Friant *et al*., 2022), wildlife population decline (Coals *et al*., 2024), and the alteration of vital ecological processes (Tyack *et al*., 2022). Additionally, people may not really be helped by these untested practices (i.e may not be treated against their diseases/ailments). To protect biodiversity, human health, and animal welfare, the belief that wild animals possess medicinal and spiritual healing properties should be discouraged. Since wild meat also drives zootherapy, ongoing efforts at monitoring and advocating against the urban wild meat trade should be intensified. Public education and risk communication, with an emphasis on the negative impacts of these practices, may be beneficial in discouraging both practitioners and their human patients.

With many people in low- and middle-income countries utilizing traditional plants for medicinal purposes (Oreagba *et al*., 2011; Iyiola *et al*., 2023), the potential of indigenous flora as viable alternatives to animal product-based treatments can be advocated, provided that it is proven to be effective, sustainable, and does not endanger plant species. This approach has been explored by some developing countries. For example, South Africa introduced conservation-friendly alternatives, such as the promotion of plant-based substitutes for perceived medicinal animal species (Green *et al*., 2022). Chinese government regulations on traditional Chinese medicines have led to the development of synthetic alternatives to products such as bile, thus reducing reliance on wild populations (Liu *et al*., 2024). Hence, promoting research on scientifically validated sustainable plant-based and synthetic healthcare products may be essential to reduce dependence on wildlife-derived products. The growing use of domestic animal products as alternatives to wildlife-derived products should also be discouraged, as this also affects animal welfare (Coals *et al*., 2024). Implementing stricter regulatory frameworks and robust enforcement mechanisms is crucial to curtail these practices to safeguard biodiversity and public health.

### 4.1. Limitations of the study

This study had a small sample size of questionnaire respondents (n=31) and focused group discussions (n=4), which were determined by the number of practitioners. No zootherapy patients provided consent to participate in the study. We acknowledge that small sample sizes result in a larger margin of error, which may impact the accuracy of our conclusions. Additionally, our reliance on self-reported data introduced potential biases, as respondents may have underreported or exaggerated certain aspects of their practices owing to cultural sensitivity and other concerns.

## Supporting information

PIS

ICF

Questionnaire

FGD Guide

## Competing interest statement

The authors declare that they have no known competing financial interests or personal relationships that could have appeared to influence the work reported in this paper.

## Funding statement

This research was supported by the International Alliance against Health Risks in Wildlife Trade, through funding by the Deutsche Gesellschaft fur Internationale Zusammenarbeit -GIZ (Grant ID: 812799425).

## Data Availability

All data produced in the present work are contained in the manuscript

## Acknowledgements

The authors wish to acknowledge Drs Olusola Ogunsanya, Ayotunde Sijuwola, Victor Assi, Ebenezer Ogundana, Oluwatobi Adedokun, Prince Offiong, Alhaji Olono, Femi Saibu, John Fadele, Cedric Mbitkebeyo and Hafeez Akinniyi whose queries and insights further shaped this research. Special thanks to the Institute of Genomics and Global Health (IGH), Redeemers University, Ede, Nigeria; Wildlife Ecology and Conservation Group of the Wageningen University & Research (WUR), Netherlands; and the One Health Center for Africa (OHRECA), International Livestock Research Institute (ILRI), Kenya, for their technical support and supervision. We also appreciate all participants across the study sites for their understanding and acceptance to participate in this study.

## Supporting information

S1 – Project information sheet (PIS) S2 – Questionnaire

S3 – Informed consent form (ICF)

S3 - Focus group discussion (FGD) Guide

## Notes

### Competing Interest Statement

The authors have declared no competing interest.

### Author Declarations

The National Human Research Ethics Committee of Nigeria (NHREC) gave ethical approval for this work

## References

1. Adebowale TK, Ijose OA, Ibiyomi BB, Akintunde OO, Oduntan OO, Osunsina IOO and Shobowale AA, 2025. Utilization of fauna resources for therapeutic purposes as a barrier to species justice advocacy in Nigeria. Front. Conserv. Sci. 6:1551597. Doi: 10.3389/fcosc.2025.1551597

2. Ademiluka, S.O., 2024. Issues of doctrine and reality in Christian attitude towards traditional medicine in Nigeria. In die Skriflig, 58(1), 3047. 10.4102/ids.v58i1.3047

3. Agusi, E.R., Allendorf, V., Eze, E.A., Asala, O., Shittu, I., Dietze, K., Busch, F., Globig, A. and Meseko, C.A., 2022. SARS-CoV-2 at the human–animal interface: implication for global public health from an African perspective. Viruses, 14(11), 2473.

4. Ahmad, S., Akram, M., Riaz, M., Munir, N., Mahmood Tahir, I., Anwar, H., Zahid, R., Daniyal, M., Jabeen, F., Ashraf, E. and Sarwar, G., 2023. Zootherapy as traditional therapeutic strategy in the Cholistan desert of BahawalpurDPakistan. Veterinary Medicine and Science, 9(4), 1861–1868.

5. Aina, O., Gautam, L., Simkhada, P. and Hall, S., 2020. Prevalence, determinants and knowledge about herbal medicine and nonhospital utilization in southwest Nigeria: a cross-sectional study. BMJ Open, 10(9), 040769.

6. Ajayi OL, Antia RE, Ojo OE, Awoyomi OJ, Oyinlola LA, Ojebiyi OG. Prevalence and renal pathology of pathogenic *Leptospira* spp. in wildlife in Abeokuta, Ogun State, Nigeria. Onderstepoort J Vet Res. 2017 Mar 24;84(1):e1–e9. doi: 10.4102/ojvr.v84i1.1210. PMID: 28397515; PMCID: PMC6238786.

7. Akpa-Inyang, F. and Chima, S.C., 2023. Traditional Health Care Practitioners’ Perspectives on Applying Informed Consent During African Traditional Medical Practice in Akwa Ibom State, Nigeria: A Cross-Sectional Qualitative Study. Journal of Integrative and Complementary Medicine, 29(6-7), 361–371.

8. Akpan, S.N., van Hooft, P., Happi, A.N., Buij, R., van Langevelde, F., Cook, E.A., Hassell, J.M., Zimmerman, D.M., Masudi, S.P., Happi, C.T. and Thomas, L.F., 2025a. Structure, conservation and health implications of urban wild meat value chains: A case study of Lagos, Nigeria. One Health, 20: 100992.

9. Akpan, S.N., Cook, E.A., Van Langevelde, F., Van Hooft, P, Zimmerman, D.M., Buij, R., Hassell, J.M., Masudi, S.P., Happi, C.T., Happi, A.N. & Thomas, L.F., 2025b. Hygiene Knowledge and Practices in the Lagos Wild Meat Value Chain: Cultural Influences, Regulatory Gaps, and Infrastructure Needs. MedRxiv, 2025-02.

10. Akunna, G.G., Lucyann, C.A. and Saalu, L.C., 2023. Rooted in tradition, thriving in the present: The future and sustainability of herbal medicine in Nigeria’s healthcare landscape. Journal of Innovations in Medical Research, 2(11), 28–40.

11. Alhaidar A., Abdel Gader A.G., Mousa S.A. The antiplatelet activity of camel urine. Journal of Alternative and Complementary Medicine, 2011;17(9):803–808. doi: 10.1089/acm.2010.0473.

12. Al-Yousef N., Gaafar A., Al-Otaibi B., Al-Jammaz I., Al-Hussein K., Aboussekhra A. Camel urine components display anticancer properties in vitro. Journal of Ethnopharmacology, 2012;143(3):819–825. doi: 10.1016/j.jep.2012.07.042.

13. Alves, R.R. and Rosa, I.L., 2005. Why study the use of animal products in traditional medicines? Journal of Ethnobiology and Ethnomedicine, 1, 1–5.

14. Alves, R.R. and Alves, H.N. 2011. The faunal drugstore: Animal-based remedies used in traditional medicines in Latin America. Journal of Ethnobiology and Ethnomedicine 7:9. 10.1186/1746-4269-7-9

15. Ammam I, Brunet CD, Boukenaoui-Ferrouk N, Peyroux J, Berthier S, Boutonnat J, Rahal K, Bitam I, Maurin M. Francisella tularensis PCR detection in Cape hares (Lepus capensis) and wild rabbits (Oryctolagus cuniculus) in Algeria. Sci Rep. 2022 Dec 12;12(1):21451. doi: 10.1038/s41598-022-25188-0. PMID: 36509808; PMCID: PMC9743112.

16. Asaaga, F.A., Tomude, E.S., Rahman, M., Shakeer, I., Ghotge, N.S., Burthe, S.J., Schäfer, S.M., Vanak, A.T., Purse, B.V. and Hoti, S.L., 2024. What is the state of the art on traditional medicine interventions for zoonotic diseases in the Indian subcontinent? A scoping review of the peer-reviewed evidence base. BMC Complementary Medicine and Therapies, 24(1), 249.

17. Awoyemi, SM., Thomas-Walters, L., Anthony BP., Vyas, D., Buij, R., Amusa, TO, 2022. Culture and the illegal trade in vultures in southwestern Nigeria: conundrums and recommendations. Vulture News, 83. Doi: 10.4314/vulnew.v83i1.2

18. Ayinmode A, Adedokun A, Aina A, Taiwo V. The zoonotic implications of pentastomiasis in the royal python (python regius). Ghana Med J. 2010 Sep;44(3):115–8. doi: 10.4314/gmj.v44i3.68895. PMID: 21327016; PMCID: PMC2996842.

19. Bachand N, Ravel A, Onanga R, Arsenault J, Gonzalez JP. Public health significance of zoonotic bacterial pathogens from bushmeat sold in urban markets of Gabon, Central Africa. J Wildl Dis. 2012 Jul;48(3):785–9. doi: 10.7589/0090-3558-48.3.785. PMID: 22740547.

20. Bagwana, P., 2015. Indigenous knowledge of traditional medicine: answering the question of knowledge acquisition and transmission among the traditional health practitioners in Uganda. Antropoloji, (30), 13-32.

21. Braun V. & Clarke V. (2006). Using thematic analysis in psychology. Qualitative Research in Psychology, 3(2), 77–101. 10.1191/1478088706qp063oa

22. Bruce HL, Barrow PA, Rycroft AN. Zoonotic potential of *Salmonella enterica* carried by pet tortoises. Vet Rec. 2018 Feb 3;182(5):141. doi: 10.1136/vr.104457. Epub 2017 Dec 7. PMID: 29217765.

23. Busse HJ, Huptas C, Baumgardt S, Loncaric I, Spergser J, Scherer S, Wenning M, Kämpfer P. Proposal of Lysobacter pythonis sp. nov. isolated from royal pythons (Python regius). Syst Appl Microbiol. 2019 May;42(3):326–333. doi: 10.1016/j.syapm.2019.02.002. Epub 2019 Feb 15. PMID: 30826139.

24. Callaway Z, Thomas A, Melrose W, Buttner P, Speare R. Salmonella Virchow and Salmonella Weltevreden in a random survey of the Asian house gecko, Hemidactylus frenatus, in houses in northern Australia. Vector Borne Zoonotic Dis. 2011 Jun;11(6):621–5. doi: 10.1089/vbz.2010.0015. Epub 2010 Oct 25. PMID: 20973656.

25. Caruso S, Edwards SJ. Recently Emerged Novel Henipa-like Viruses: Shining a Spotlight on the Shrew. Viruses. 2023 Dec 11;15(12):2407. doi: 10.3390/v15122407. PMID: 38140648; PMCID: PMC10747904.

26. Challender, D.W., Heinrich, S., Shepherd, C.R. and Katsis, L.K., 2020. International trade and trafficking in pangolins, 1900–2019. In pangolins, Academic Press, pp. 259–276.

27. Cléry-Barraud, C., Gaubert, A., Masson, P., & Vidal, D. 2004. Combined effects of high hydrostatic pressure and temperature for inactivation of *Bacillus anthracis* spores. Applied and environmental microbiology, 70(1), 635–637. 10.1128/AEM.70.1.635-637.2004

28. Coals, P.G., Mbongwa, N.S., Naude, V.N. and Williams, V.L., 2022. Contemporary cultural trade of lion body parts. Animals, 12(22), 3169.

29. Coals PGR, Williams VL, Benítez G, Chassagne F, Leonti M. 2024. Ethnopharmacology, ethnomedicine, and wildlife conservation. J Ethnopharm. 333:118399. 10.1016/j.jep.2024.118399

30. Coppola F, Cilia G, Bertelloni F, Casini L, D’Addio E, Fratini F, Cerri D, Felicioli A. Crested Porcupine (Hystrix cristata L.): A New Potential Host for Pathogenic Leptospira Among Semi-Fossorial Mammals. Comp Immunol Microbiol Infect Dis. 2020 Jun;70:101472. doi: 10.1016/j.cimid.2020.101472. Epub 2020 Mar 19. PMID: 32208192.

31. Dutch Wildlife Health Centre. Zoonotic diseases in hares. Apr 2015. Available online at: https://dwhc.nl/en/2015/04/zoonotic-diseases-in-hares/ (Accessed on 22/102025)

32. Ebani VV. Domestic reptiles as source of zoonotic bacteria: A mini review. Asian Pac J Trop Med. 2017 Aug;10(8):723–728. doi: 10.1016/j.apjtm.2017.07.020. Epub 2017 Aug 23. PMID: 28942820.

33. Elendu, C., 2024. The evolution of ancient healing practices: From shamanism to Hippocratic medicine: A review. Medicine, 103(28), 39005.

34. Esteves SB, Gaeta NC, Batista JMN, Dias RA, Heinemann MB. Leptospira sp. infection in bats: A systematic review and meta-analysis. Transbound Emerg Dis. 2022 Sep;69(5):e2456–e2473. doi: 10.1111/tbed.14589. Epub 2022 May 30. PMID: 35533065.

35. Ernst E. How the public is being misled about complementary/alternative medicine. J R Soc Med. 2008 Nov; 101(11):528–30. doi: 10.1258/jrsm.2008.080233

36. Fourchault, L., Lamane, A., Nguinwa Mbakop, D.R., Saliu, G.T., Gryseels, S., Verheyen, E. and Kreppel, K., 2025. Public health risks of traditional zootherapeutic practices in Africa, One Health, 21:101178. 10.1016/j.onehlt.2025.101178

37. Friant, S., Bonwitt, J., Ayambem, W.A., Ifebueme, N.M., Alobi, A.O., Otukpa, O.M., Bennett, A.J., Shea, C., Rothman, J.M., Goldberg, T.L. and Jacka, J.K., 2022. Zootherapy as a potential pathway for zoonotic spillover: a mixed-methods study of the use of animal products in medicinal and cultural practices in Nigeria. One Health Outlook, 4(1), 5–12.

38. Gao, H., Dou, H., Wei, S., Sun, S., Zhang, Y. and Hua, Y., 2022. Local chronicles reveal the effect of anthropogenic and climatic impacts on local extinctions of Chinese pangolins (*Manis pentadactyla*) in mainland China. Ecology and Evolution, 12(10), 9388.

39. Green, J., Hankinson, P., de Waal, L., Coulthard, E., Norrey, J., Megson, D. and D’Cruze, N., 2022. Wildlife trade for belief-based use: Insights from traditional healers in South Africa. Frontiers in Ecology and Evolution, 10, 906398.

40. Grigo, J., 2022. Traditional African Medicine as Living Cultural Heritage: Conditions and Politics of Knowledge Transfer. Senri Ethnological Studies, 109, 101–124.

41. Gurumyen, B.D., Akanle, O., Yikwabs, Y.P. and Nomishan, T.S., 2020. Zootherapy: The use of dog meat for traditional African medicine in Kanke local government area, Plateau state, Nigeria. Journal of Tourism and Heritage Studies, 10.33281/JTHS20129.2020.2.1.hal-03548339.

42. Hahn RA, Truman BI. Education improves public health and promotes health equity. Int J Health Serv. 2015;45(4):657–78. doi: 10.1177/0020731415585986. Epub 2015 May 19. PMID: 25995305; PMCID: PMC4691207.

43. Happi AN, Olumade TJ, Ogunsanya OA, Sijuwola AE, Ogunleye SC, Oguzie JU, Nwofoke C, Ugwu CA, Okoro SJ, Otuh PI, Ngele LN, Ojo OO, Adelabu A, Adeleye RF, Oyejide NE, Njaka CS, Heeney JL, Happi CT. Increased Prevalence of Lassa Fever Virus-Positive Rodents and Diversity of Infected Species Found during Human Lassa Fever Epidemics in Nigeria. Microbiol Spectr. 2022 Aug 31;10(4):e0036622. doi: 10.1128/spectrum.00366-22. Epub 2022 Aug 1. PMID: 35913205; PMCID: PMC9430508.

44. Haq, Z., Nazir, J., Manzoor, T., Saleem, A., Hamadani, H., Khan, A.A., Bhat, S.S., Jha, P. and Ahmad, S.M., 2024. Zoonotic spillover and viral mutations from low and middle-income countries: improving prevention strategies and bridging policy gaps. PeerJ, 12, 17394. 10.7717/peerj.17394

45. Hogerwerf L, Roof I, de Jong MJK, Dijkstra F, van der Hoek W. Animal sources for zoonotic transmission of psittacosis: a systematic review. BMC Infect Dis. 2020 Mar 4;20(1):192. doi: 10.1186/s12879-020-4918-y. PMID: 32131753; PMCID: PMC7057575.

46. Inatimi, S.A., Popoola, O.M., Yarkwan, B., Iyiola, A.O. and Izah, S.C., 2022. Therapeutic potentials of wildlife resources and options for conservation. In Biodiversity in Africa: potentials, threats and conservation, Singapore: Springer Nature, pp. 143–174.

47. International Union for Conservation of Nature (2023). Red List of Threatened Species. Available at: www.iucnredlist.org. Accessed 24 February 2025.

48. Iyiola, A.O., Adegoke Wahab, M.K., 2023. Herbal Medicine Methods and Practices in Nigeria. In: Izah, S.C., Ogwu, M.C., Akram, M. (eds) Herbal Medicine Phytochemistry. Reference Series in Phytochemistry. Springer, Cham. 10.1007/978-3-031-21973-3_47-1

49. Jegede HO, Daodu OB, Adesiji YO and Shafi AA. 2016. Prevalence of Salmonella species isolated from the African Rock Python. J Wildlife Res. 4(2):32–34

50. Jiang X, Fan Z, Li S, Yin H. A Review on Zoonotic Pathogens Associated with Non-Human Primates: Understanding the Potential Threats to Humans. Microorganisms. 2023 Jan 18;11(2):246. doi: 10.3390/microorganisms11020246. PMID: 36838210; PMCID: PMC9964884.

51. Kendie, F.A., Mekuriaw, S.A. and Dagnew, M.A., 2018. Ethnozoological study of traditional medicinal appreciation of animals and their products among the indigenous people of Metema Woreda, North-Western Ethiopia. Journal of Ethnobiology and Ethnomedicine, 14, 1–12.

52. Koski L, DeBess E, Rosen HE, Reporter R, Waltz T, Leeper M, Concepcion Acevedo J, Karpíšková R, McCormick J, Gelbicova T, Morningstar-Shaw B, Nichols M, Leman RF. An investigation of Salmonella Fluntern illnesses linked to leopard geckos-United States, 2018. Zoonoses Public Health. 2019 Dec;66(8):974-977. doi: 10.1111/zph.12647. Epub 2019 Sep 11. PMID: 31512375.

53. Kuralkar, P. and Kuralkar, S.V., 2021. Role of herbal products in animal production–An updated review. Journal of Ethnopharmacology, 278, 114246.

54. Lam TT, Jia N, Zhang YW, Shum MH, Jiang JF, Zhu HC, Tong YG, Shi YX, Ni XB, Liao YS, Li WJ, Jiang BG, Wei W, Yuan TT, Zheng K, Cui XM, Li J, Pei GQ, Qiang X, Cheung WY, Li LF, Sun FF, Qin S, Huang JC, Leung GM, Holmes EC, Hu YL, Guan Y, Cao WC. Identifying SARS-CoV-2-related coronaviruses in Malayan pangolins. Nature. 2020 Jul;583(7815):282-285. doi: 10.1038/s41586-020-2169-0. Epub 2020 Mar 26. PMID: 32218527.

55. Laughlin, C.D. and Rock, A.J., 2014. What can we learn from shamans’ dreaming? A cross-cultural exploration. Dreaming, 24(4), 233.

56. Lee, T.M., Sigouin, A., Pinedo-Vasquez, M. and Nasi, R., 2020. The harvest of tropical wildlife for bushmeat and traditional medicine. Annual Review of Environment and Resources, 45(1), 145–170.

57. Lemhadri, A., Benali, T., Tekalign, W. and Wendimu, A., 2024. Zootherapeutic uses of animals and their parts: An important element of the traditional knowledge of the Safi province, Morocco. Heliyon, 10(22), 23–30.

58. Liu P, Jiang JZ, Wan XF, Hua Y, Li L, Zhou J, Wang X, Hou F, Chen J, Zou J, Chen J. Are pangolins the intermediate host of the 2019 novel coronavirus (SARS-CoV-2)? PLoS Pathog. 2020 May 14;16(5):e1008421. doi: 10.1371/journal.ppat.1008421.

59. Liu, X., Li, S., Feng, Y., Chen, X., Ma, Y., Xiao, H., Zhao, Y., Liu, S., Zheng, G., Yang, X. and Wu, F., 2024. Traditional knowledge of animal-derived medicines used by Gelao community in Northern Guizhou, China. Journal of Ethnobiology and Ethnomedicine, 20(1), 31.

60. Magouras, I., Brookes, V.J., Jori, F., Martin, A., Pfeiffer, D.U. and Dürr, S., 2020. Emerging zoonotic diseases: should we rethink the animal–human interface? Frontiers in Veterinary Science, 7, 582743.

61. Masudi SP, Hassell J, Cook EA, van Hooft P, van Langevelde F, Buij R, Otiende MY, Ochieng JW, Santangeli A, Happi A, Akpan SN, Thomas LF. Limited knowledge of health risks along the illegal wild meat value chain in the Nairobi Metropolitan Area (NMA). PLoS One. 2025 Mar 26;20(3):e0316596. doi: 10.1371/journal.pone.0316596. PMID: 40138327; PMCID: PMC11940438.

62. Mendoza-Roldan JA, Mendoza-Roldan MA, Otranto D. Reptile vector-borne diseases of zoonotic concern. Int J Parasitol Parasites Wildl. 2021 Apr 22;15:132–142. doi: 10.1016/j.ijppaw.2021.04.007. PMID: 34026483; PMCID: PMC8121771.

63. Mishra, B., Akhila, M.V., Thomas, A., Benny, B. and Assainar, H., 2020. Formulated Therapeutic Products of Animal Fats and Oils: Future Prospects of Zootherapy. Int J Pharm Invest, 10(2), 1–10.

64. Mitchell, J., 2023. Antimicrobial resistance (AMR) as a form of human–wildlife conflict: Why and how nondomesticated species should be incorporated into AMR guidance. Ecology and Evolution, 13(9), 10421.

65. Mlangeni LN, Ramatla T, Lekota KE, Price C, Thekisoe O, Weldon C. Occurrence, Antimicrobial Resistance, and Virulence Profiles of *Salmonella* Serovars Isolated from Wild Reptiles in South Africa. Int J Microbiol. 2024 Jan 5;2024:5213895. doi: 10.1155/2024/5213895. PMID: 38222969; PMCID: PMC10787053.

66. Mukherjee, S., Gomes, A. and Dasgupta, S.C., 2017. Zoo therapeutic uses of snake body parts in folk & traditional medicine. Journal of Zoological Research, 1(1), 1–9.

67. Nanda, S., 2023. Integrating traditional and contemporary systems for health and well-being. Annals of neurosciences, 30(2), 77–78.

68. Neiffer D, Hewlett J, Buss P, Rossouw L, Hausler G, deKlerk-Lorist LM, Roos E, Olea-Popelka F, Lubisi B, Heath L, Miller M. Antibody prevalence to African swine fever virus, Mycobacterium bovis, Foot-and-mouth disease virus, Rift valley fever virus, Influenza A virus, and Brucella and Leptospira spp. In free-ranging warthog (*Phacochoerus africanus*) populations in South Africa. J Wildl Dis. 2021 Jan 6;57(1):60–70. doi: 10.7589/JWD-D-20-00011. PMID: 33635986.

69. Nevarez J. CROCODILIANS. Manual of Exotic Pet Practice. 2009:112–35. doi: 10.1016/B978-141600119-5.50009-3. Epub 2009 Nov 30. PMCID: PMC7152205.

70. Nieman, W.A., Leslie, A.J. and Wilkinson, A., 2019. Traditional medicinal animal use by Xhosa and Sotho communities in the Western Cape Province, South Africa. Journal of Ethnobiology and Ethnomedicine, 15, 1–14.

71. Ogunshe, A., Fawole, A. and Ajayi, V., 2010. Microbial evaluation and public health implications of urine as an alternative therapy in clinical pediatric cases: health implications of urine therapy. Pan African Medical Journal, 5(1).

72. Oreagba, I.A., Oshikoya, K.A. and Amachree, M., 2011. Herbal medicine use among urban residents in Lagos, Nigeria. BMC Complementary and Alternative medicine, 11, 1–8.

73. Patrick, P.G. and Singkam, A.R., 2024. Biodiversity conservation, human–animal interactions, and zootherapy in ecological knowledge of Indonesian Healers. Conservation Biology, 38(4), 14278.

74. Panti-May JA, DE Andrade RRC, Gurubel-González Y, Palomo-Arjona E, Sodá-Tamayo L, Meza-Sulú J, Ramírez-Sierra M, Dumonteil E, Vidal-Martínez VM, Machaín-Williams C, DE Oliveira D, Reis MG, Torres-Castro MA, Robles MR, Hernández-Betancourt SF, Costa F. A survey of zoonotic pathogens carried by house mouse and black rat populations in Yucatan, Mexico. Epidemiol Infect. 2017 Aug;145(11):2287–2295. doi: 10.1017/S0950268817001352. Epub 2017 Jul 10. PMID: 28689507; PMCID: PMC6231242.

75. Plaza PI, Blanco G, Madariaga MJ, Boeri E, Teijeiro ML, Bianco G, Lambertucci SA. Scavenger birds exploiting rubbish dumps: Pathogens at the gates. Transbound Emerg Dis. 2019 Mar;66(2):873–881. doi: 10.1111/tbed.13097. Epub 2018 Dec 28. PMID: 30548806.

76. Račka K, Bártová E, Hamidović A, Plault N, Kočišová A, Camacho G, Mercier A, Halajian A. First detection of *Toxoplasma gondii* Africa 4 lineage in a population of carnivores from South Africa. Front Cell Infect Microbiol. 2024 Jan 30;14:1274577. doi: 10.3389/fcimb.2024.1274577. PMID: 38352059; PMCID: PMC10861644.

77. Rajasekharan, S., Nair, V.T., Navas, M., James, T.C. and Murugan, K., 2023. Traditional knowledge and its sustainable utilization. Conservation and Sustainable Utilization of Bioresources, 597-657.

78. Rush, E.R., Dale, E. and Aguirre, A.A., 2021. Illegal wildlife trade and emerging infectious diseases: Pervasive impacts to species, ecosystems and human health. Animals, 11(6), 1821.

79. Saidu, Y. & Buij, R. (2013) Traditional medicine trade in vulture parts in northern Nigeria. Vulture News, 65, 4–14.

80. Santangeli, A., Lambertucci, S.A., Margalida, A., Carucci, T., Botha, A., Whitehouse-Tedd, K. and Cancellario, T., 2024. The global contribution of vultures towards ecosystem services and sustainability: An experts’ perspective. Iscience, 27(6), 109925.

81. Schmidt K, Pittler MH, Ernst E. Bias in alternative medicine is still rife but is diminishing. BMJ. 2001 Nov 3;323(7320):1071. PMID: 11691776; PMCID: PMC1121565.

82. Schneeberger, K., Voigt, C.C. (2016). Zoonotic Viruses and Conservation of Bats. In: Voigt, C., Kingston, T. (eds) Bats in the Anthropocene: Conservation of Bats in a Changing World. Springer, Cham. 10.1007/978-3-319-25220-9_10

83. Simulundu, E., Ndashe, K., Chambaro, H. M., Squarre, D., Reilly, P., Chitanga, S.…Sawa, H. (2020). West Nile Virus in Farmed Crocodiles, Zambia, 2019. Emerging Infectious Diseases, 26(4), 811–814. 10.3201/eid2604.190954.

84. Soewu, D. A. 2008. Wild animals in ethnozoological practices among the Yorubas of southwestern Nigeria and the implications for biodiversity conservation. African Journal of Agricultural Research, 3: 421–427.

85. Soewu, D.A., 2012. Zootherapy and biodiversity conservation in Nigeria. In Animals in Traditional Folk Medicine: Implications for Conservation. Berlin, Heidelberg: Springer Berlin Heidelberg, pp. 347–365.

86. Souto, W.M., Mourao, J.S., Barboza, R.R.D. and Alves, R.R., 2011. Parallels between zootherapeutic practices in ethnoveterinary and human complementary medicine in northeastern Brazil. Journal of ethnopharmacology, 134(3), 753–767.

87. Spinks AT, Dunstan RH, Harrison T, Coombes P, Kuczera G. Thermal inactivation of water-borne pathogenic and indicator bacteria at subboiling temperatures. Water Res. 2006 Mar;40(6):1326–32. doi: 10.1016/j.watres.2006.01.032. PMID: 16524613.

88. Suwanpakdee S, Bhusri B, Saechin A, Mongkolphan C, Tangsudjai S, Suksai P, Kaewchot S, Sariwongchan R, Sereerak P, Sariya L. Potential Zoonotic Infections Transmitted by Free-Ranging Macaques in Human-Monkey Conflict Areas in Thailand. Zoonoses Public Health. 2025 Jun;72(4):349–358. doi: 10.1111/zph.13211. Epub 2025 Jan 23. PMID: 39853967; PMCID: PMC12016002.

89. Swift, B.M., Bennett, M., Waller, K., Dodd, C., Murray, A., Gomes, R.L., Humphreys, B., Hobman, J.L., Jones, M.A., Whitlock, S.E. and Mitchell, L.J., 2019. Anthropogenic environmental drivers of antimicrobial resistance in wildlife. Science of the Total Environment, 649, 12–20.

90. Tajudeen, Y.A., Oladunjoye, I.O., Bajinka, O. and Oladipo, H.J., 2022. Zoonotic spillover in an era of rapid deforestation of tropical areas and unprecedented wildlife trafficking: into the wild. Challenges, 13(2), 41.

91. Tan, M., Otake, Y., Tamming, T., Akuredusenge, V., Uwinama, B. and Hagenimana, F., 2021. Local experience of using traditional medicine in northern Rwanda: a qualitative study. BMC Complementary Medicine and Therapies, 21, 1–11.

92. Tawiah-Mensah CNL, Ladzekpo D, Addo SO. Bushmeat Consumption and the Risk of Zoonotic Tick-Borne Pathogen Infections in Ghana: An Increasing Risk to Public Health. Public Health Chall. 2025 Jul 30;4(3):e70096. doi: 10.1002/puh2.70096. PMID: 40741378; PMCID: PMC12309406.

93. Tijani, M., 2024. History and Challenges Facing Traditional Healing Practices in Nigeria. Available at: https://wap.org.ng/read/traditional-healing-practices-in-nigeria/ (accessed 24 February 2025).

94. Tran, T. and Xie, S., 2024. Mitigating wildlife spillover in the clinical setting: how physicians and veterinarians can help prevent future disease outbreaks. AJPM Focus, 3(2), 100193.

95. Traversa D, Iorio R, Otranto D, Modrý D, Slapeta J. Cryptosporidium from tortoises: Genetic characterisation, phylogeny and zoonotic implications. Mol Cell Probes. 2008 Apr;22(2):122–8. doi: 10.1016/j.mcp.2007.11.001. Epub 2007 Nov 23. PMID: 18234467.

96. Tumelty, L., Fa, J.E., Coad, L., Friant, S., Mbane, J., Kamogne, C.T., Tata, C.Y. and Ickowitz, A., 2023. A systematic mapping review of links between handling wild meat and zoonotic diseases. One Health, 17, 100637.

97. Tyack PL, Thomas L, Costa DP, Hall AJ, Harris CM, Harwood J, Kraus SD, Miller PJO, Moore M, Photopoulou T, Pirotta E, Rolland RM, Schwacke LH, Simmons SE, Southall BL. Managing the effects of multiple stressors on wildlife populations in their ecosystems: developing a cumulative risk approach. Proc Biol Sci. 2022 Nov 30;289(1987):20222058. doi: 10.1098/rspb.2022.2058. Epub 2022 Nov 30. PMID: 36448280; PMCID: PMC9709579.

98. Wilkins, P. A. 2014. Botulism. Chapter 43 - Equine Infectious Diseases (2^nd^ Edition), (Ed. Debra C. Sellon, Maureen T. Long). W.B. Saunders, 2014; 364-367.e2. 10.1016/B978-1-4557-0891-8.00043-9.

99. Williams, V.L., Cunningham, A.B., Kemp, A.C. & Bruyns, R.K. (2014). Risks to birds traded for African traditional medicine: a quantitative assessment. PLoS ONE, 9 (8): e105397.

100. Zhao W, Zhang Y, Li J, Ren G, Qiang Y, Wang Y, Lai X, Lei S, Liu R, Chen Y, Huang H, Li W, Lu G, Tan F. Prevalence and distribution of subtypes of Blastocystis in Asiatic brush-tailed porcupines (Atherurus macrourus), bamboo rats (Rhizomys pruinosus), and masked palm civets (Paguma larvata) farmed in Hainan, China. Parasite. 2023;30:45. doi: 10.1051/parasite/2023048. Epub 2023 Nov 2. PMID: 37921619; PMCID: PMC10624160.

101. Zhou, P., Yang, X.L., Wang, X.G., Hu, B., Zhang, L., Zhang, W., Si, H.R., Zhu, Y., Li, B., Huang, C.L. and Chen, H.D., 2020. A pneumonia outbreak associated with a new coronavirus of probable bat origin. Nature, 579(7798): 270–273.

