## Supplementary material for "Use of animal-derived products for medicinal and belief-based purposes in urban cities of southwestern Nigeria: a One Health perspective": PIS

**Principal Investigators:**

Anise Happi, PhD.

Phone (Nigeria): (+234) 802 338 3683

Christian Happi, PhD.

Phone (Nigeria): (+234) 802 338 3684

**Field Coordinator:**

Samuel Akpan, MVPH

Phone (Nigeria): (+234) 7035332772

**Performance Site:**

Osun, Ogun, Ondo Oyo, and Lagos states, Nigeria.

**Introduction**

We are researchers from the African Centre of Excellence for Genomics of Infectious Diseases (ACEGID), Redeemers University, Ede, Osun State. You have been invited to participate in a research study to find the source and risk factors for transmission of pathogens from wildlife to humans through bushmeat. You have been selected because what we are searching for can be found in the wild animals and bushmeat products you regularly come in contact with.

**Why is this study being done?**

Pathogens are tiny germs that we cannot see with our naked eyes, but can cause mild to serious diseases. Most of the ones that cause diseases in humans can come from wild animals who themselves may or may not be affected. It can also lead to the disappearance of some wildlife from our forests, due to over-use. There are various ways in which these pathogens are transmitted from to humans causing diseases. One of the possible ways is through wild meat. We are carrying out this study to answer the following questions:

- *Which species are traded in the Lagos wild meat value chain?*
- *Who are the actors, their characteristics and governance structure?*
- *Are there pathogens (germs) in the wild meat traded in Lagos?*
- *What are the practices and knowledge of actors that can make it easier for humans to contract diseases through wild meat.*
- *Who are the people most likely to be affected, and at what point are they likely to be affected?*
- *Are the germs in the bushmeat capable of causing large disease outbreaks now or in the future?*

The findings of this research will help to prevent and control the transmission of pathogens from wildlife to humans; and also help in designing solutions to protect wildlife. The isolated pathogens can be used to develop diagnostic kits which will enable early detection of possible infection in humans so to stop infectious disease from progressing. This research may also contribute to the development of policies that can help us prevent future disease outbreaks.

**What are the study procedures and what will I be asked to do?**

If you agree to be involved in this study,

- We will visit your business point or market to ask you a few questions and your responses will be recorded for analysis.
- We will ask you to fill a questionnaire (you may be assisted on this by our researchers).
- If there is fresh unprocessed or processed bushmeat, we may take samples (swabs or small tissue cutting) where possible or as necessary.
- The collected samples will be packaged and taken to our laboratory at African Centre of Excellence for Genomics of Infectious Diseases (ACEGID) to test for the presence of any pathogens of interest.
- During the study, we will obtain general information about you and some of the activities that you are involved in. We assure you that your name will not be attached to this study except the GPS coordinates and the sampling addresses generated by us for easy identification.
- The test results will be made known to the researchers (the scientists) working on the study who may contact you if there is a need for further information.
- Stored samples will have a unique number that may be linked to the GPS coordinates of the market/sales point, and may be used for further studies.
- An ethics board will review any future study by these and other researchers and determine if it is ethical to use these samples for the research based on the information provided in this informed consent.
- We will not be able to notify you directly to tell you more information about those potential future studies.
- You do not have to answer any of the questions by our researchers if you are not comfortable with them.

**What other options are there?**

The alternative to participating in the study is not to participate. Choosing to participate or not participate is voluntary and will not affect your daily activities.

**What are the risks or inconveniences of this study?**

- There are no risks associated with this study. The time taken to answer questions or fill the questionnaire may disrupt your daily activities causing inconvenience to you. We will ensure that the timing for the interview or filling of questionnaire is properly discussed with you to minimize the disruption that may arise from participating in this study.
- There may be risks to this study that are currently unforeseeable. We will do everything within our power to reduce these risks as they arise.

**What are the benefits of the study?**

There is no direct benefit to you as a result of participating in this research. However, this study will contribute to the detection of pathogens present in bushmeat, which may also be of interest to you. We also hope that your participation in this study will enable us to understand how people get sick from the pathogens, which may enable better understanding of how to preventing, or even eradicate them completely. We will communicate the general results of this study to the market association and community leaders to share with the community. If we will be taking samples that may affect the sales of your bushmeat, be assured that we will compensate accordingly by either paying for the part or whole.

**Will I receive payment for participation?**

You will not be paid for participating in this study.

**Are there costs for participation?**

There are no costs to you to participate in this study.

**Possible Commercial Products**

The samples and information collected from this study may be used for research purposes as described in this consent form. Furthermore, if you agree below, they may be stored and used for additional research in the future. You understand and agree that once you provide consent for this study, researchers at ACEGID, Redeemer’s University, Wageningen University, and other collaborators will have the right to carry out research with the samples and information collected from your market or sales point as they no longer belong to you. It is possible that some of the research carried out with the samples we are collecting or some of the information in the research databases may help researchers develop new medical tests, or new medicines to treat or prevent diseases. These tests or products may be able to help people get well or stay healthy and so they may be sold. However, Redeemer’s University and collaborators will work with the government of Nigeria and third parties to ensure that any products developed from the sample will be made available to the people of Nigeria at minimal or no costs.

**How will my personal information be protected?**

No personal information will be collected for this study. All the samples will be linked to the GPS coordinates or the assigned number to your shop or sales point. However, the following procedures will be used to protect the confidentiality of your data:

- We will keep all study records (including any codes to store or stand data) locked in a secure location.
- Research records will be labeled with a unique code. A master key that links GPS coordinates/ address and codes will be maintained in a separate and secure location for 7 years after the conclusion of the study.
- All electronic files (e.g., database, spreadsheet, etc.) containing identifiable information will be password protected. Any computer hosting such files will also have password protection to prevent access by unauthorized users.
- Only the specific members of our research staff will have access to the passwords. Data that will be shared with others will be coded as described.
- At the conclusion of this study, the we may publish our findings. However, information collected during the course of this study will be presented in summary format and your shops or sales point will not be identified in any publications or presentations.
- Sales point or area data, without any identifier may also be shared with and stored in a national data bank available to approved researchers only.
- Any master key, and other data described above will be maintained in accordance with the security procedures until destroyed by the researchers.

You should also know that the Redeemer’s University Animal Research Protection Office and the Biomedical Institutional Review Board (IRB), and other collaborator IRBs may inspect study records as part of their auditing program. However, these reviews will only focus on the researchers and not on your responses or involvement. The IRB is a group of people who review research studies to protect the rights and welfare of research participants.

**Disclosure of Potential Conflicts of Interest**

Some of the researchers working in this study are also healthcare providers. They are interested in the knowledge to be gained from this study and in your well-being. Investigators may obtain salary or other financial support for conducting the research. You are under no obligation to participate in the research study offered to you.

**Can I stop being in the study and what are my rights?**

You do not have to be in this study if you do not want to. If you agree to be in the study, but later change your mind, you may drop out at any time. There are no penalties or consequences of any kind if you decide that you do not want to participate. However, because your responses and the samples here are animal samples which are not tied to any individual, the samples and data will continue to be used for research. Nonetheless, we will ensure that everything linking the samples to you to the extent that we are still able to identify the household will be destroyed.

**Who do I contact if I have questions about the study?**

Take as much time as you need before you decide to participate in this study. We will be happy to answer any questions you have about this study. If you have any questions about the study or any problems with the study you can contact Anise Happi (Phone: +234 802 338 3683;), Akpan Samuel (Phone: +234 703 533 2772;

**Who is conducting this study?**

Investigators at Redeemers University are conducting this study. The principal investigator who oversees Animal subject research is Dr Happi Anise at ACEGID, Ede, Osun State, Nigeria.

**YOU WILL BE GIVEN A COPY OF THIS SHEET**

*Thank you for considering to take part in this study. The investigator must explain this study to you before you agree to take part. If you have any questions arising from the information given to you, please ask the investigator conducting the research before you decide whether to take part or not. You will be given a copy of this form to keep and refer to at any time.*

**Documentation of Consent:**

I have read or received information from this form and decided that I will participate in the research project described in this form. Its general purposes, the particulars of involvement and possible risks and inconveniences have been explained to my satisfaction. I understand that I do not have to participate in the study and can withdraw at any time. My signature also indicates that I have received a copy of this consent form. Each page of the consent form is initiated by me and the study staff, to indicate that the study staff has reviewed all of the pages with me.

By ticking each box, you are consenting to the elements of this study. It will be assumed that unticked box means that you DO NOT consent to that part of the study, and this may make you ineligible to participate in the study.

I agree to allow the study staff initials on each page of the consent form.

There might be other research questions that investigators could study in the future with the help of samples like the one collected from the animals.

I agree to fill the questionnaire, or take part in interview and discussions.

I understand I will not be re-contacted about future potential use.

I understand that my participation is voluntary and that I do not have to take part if I do not want to. I understand that I am free to withdraw from this study if I change my mind.

____________________________________________	_____________

Subject name and signature	Date

____________________________________________	_____________

Parent/Legally Authorized Representative (if applicable)	Date

I am unable to read but this consent document has been read and explained to me by ___________________ (name of reader). I therefore volunteer to participate in this research.

____________________________________________	_____________

Subject	name and signature	Date

____________________________________________	_____________

Witness	Date

### Signature of Investigator or Responsible Individual:

To the best of my ability, I __________________________(name of investigator / person obtaining consent) have explained and discussed the full contents of the study, including all information contained in this consent form, and I have answered all questions from the research subjects and those of his/her parent(s) or legal guardian.

____________________________________________	_____________

Signature	Date
