## Supplementary material for "Use of animal-derived products for medicinal and belief-based purposes in urban cities of southwestern Nigeria: a One Health perspective": Questionnaire

**Study Title:** Investigating zootherapy and wild meat practices in Lagos and South west Nigeria.

**Principal Investigators:** Anise Happi & Christian Happi.

**Field Coordinator:** Samuel Akpan

**Questionnaire ID:**

**Date:**

**Area:**

Do you consent to participate in this study? Yes	No

** Kindly tick the option(s) as it applies to you.*

A. **Demographics of the participant**

*(This section is for your basic personal information. This information will help us better understand actors in the value chain).*

1. What is your age?
a) ≤18 years
b) 18-40 years
c) 40-55 years
d) ≥55 years
2. What is your gender?
a) Female
b) Male
3. What is your level of education?
a) Primary
b) Secondary
c) College
d) University
e) Informal education
f) None
4. What is your religion?
a) Muslim
b) Christian
c) Traditionalist
d) Others
B. **Value Chain Structure**

*(This section seeks to assess your role in the wild meat value chain, other actors involved, species traded, the temporal and spatial characteristics of the value chain).*

5. What is your role in the value chain?
(a) Hunter
(b) Wholesaler
(c) Processor
(d) Retailer
(e) Consumer
(f) Others (please specify)
6. For what purposes do you trade in wildmeat?
a) For income
b) For food
c) For religious purposes
d) For medicinal purposes
e) Other reasons (please specify)
7. Which animal(s) species do you trade?
8. How often do you trade wild meat?
a) every day
b) every week
c) every month
d) Occasionally
e) I don´t know
9. Which season is the busiest for your activities?
a) Rainy season
b) Dry season
c) All seasons
d) None
10. What time of the day is your business mostly active?
(a) In the Day time (6 pm-6 am)
(b) At night (6 pm-6 am)
(c) At all times (both night & day)
11. From whom do you obtain the wild meat?
a) I hunt it myself
b) Hunters
c) Wholesalers
d) Processors
e) Retailers__________________________________________________
12. To whom do you supply/sell your products?

a)	Hunters

b)	Wholesalers

c)	Processors

d)	Retailers

e) Consumers (please specify)

13. From which location do you obtain the wild meat?
(a) Within Lagos
(b) Outside Lagos (please specify)
(c) I do not know
14. Do you process wild meat in any way?
a) Yes
b) No
15. If yes to the question above, how do you process the wild meat?
a) Skinning
b) Evisceration
c) Salting
d) Smoking
e) Grilling
f) Boiling/cooking
g) Other (please specify)
C. **Zootherapy and Belief-Based Practices**

*(This section seeks to assess your practices with regards to use of wildlife products for healthcare or religious purposes, and the species used for these purposes in the value chain).*

16. Apart from eating, do you use wild animals for other purposes?
(a) Yes
(b) No
(c) I prefer not to say
17. If yes to above, for what other purposes do you use wild animals?

……………………………………………………………………………………………..

……………………………………………………………………………………………..

……………………………………………………………………………………………..

……………………………………………………………………………………………..

18. Which species and parts do you use?

……………………………………………………………………………………………..

……………………………………………………………………………………………..

19. Where do you source the wildlife or wildlife-derived products?

………………………………………………………………………………………………

20. How many years have you practiced this?
a) Less than 10 years
b) 10-20 years
c) 20-30 years
d) 30-40 years
e) 40-50 years
f) More than 50 years
21. How did you acquire the knowledge of this practice?

……………………………………………………………………………………………..

22. Do you think this practice is effective (do you get positive results)?
a) Yes
b) No
c) I don’t know
23. Do you think that this practice has any public health risks?
a) Yes
b) No
c) I don’t know
24. Do you think that this practice is can cause harm to wildlife population?
a) Yes
b) No
c) I don’t know
25. Do you think that these practices have impact on the environment?
d) Yes
e) No
f) I don’t know

**Thank you for participating in this study**
