## Supplementary material for "Use of animal-derived products for medicinal and belief-based purposes in urban cities of southwestern Nigeria: a One Health perspective": FGD Guide

**Study Title:** Health Risks of Urban Wild Meat in Lagos, Nigeria.

**[Sub-title:** Medicinal and belief-based use of wildlife products in southwest Nigeria]

[Duration: 45 minutes]

(1) What is (are) your profession(s)?
(2) Do you use wildlife products to treat human diseases and other belief-based purposes?
(3) Where are you based, and are there any possible reasons why you chose the location?
(4) Which wild animal species do you use in your practice?
(5) Kindly mention some of the practices for which you use wildlife products
(6) Do you think that the practice of using wildlife products for treatment of humans can cause harm to wildlife?
(7) **Are the commonly used species still abundant today, as in the past?**
(8) **Do you use one animal body repeatedly for treatment of more than one person?**
(9) **Do you treat the products before use? If yes, how?**
(10) **Do you think that using wildlife products for human treatments parts can lead to transmission of diseases to humans?**
(11) **Where, and from whom do you source the wildlife products**
(12) **Do you think your practice is very** effective?
(13) Is there any link between your practice and wild meat trade? Are you also involved in both?
